# Structural basis of fitness of emerging SARS-COV-2 variants and considerations for screening, testing and surveillance strategy to contain their threat

**DOI:** 10.1101/2021.01.28.21250666

**Authors:** Sk Ramiz Islam, Debasish Prusty, Soumen Kanti Manna

## Abstract

While emergence of new SAS-COV-2 variants is posing grave challenge to efforts to deal with the COVID-19 pandemic, the structural and molecular basis of their fitness remain poorly understood. We performed *in silico* analysis of structures of two most frequent SARS-COV-2 mutations, namely, N501Y and E484K, to identify plausible basis of their fitness over the original strain. The analysis suggested that the N501Y mutation is associated with strengthening of intra- as well as intermolecular H-bond in the hACE2 receptor-spike protein complex, which could result in increased affinity and, therefore, higher infectivity. While E484K mutation did not seem to directly affect the binding with hACE2 receptor, it disrupted H-bonding and salt-bridge interaction associated with binding with neutralizing antibody, which could affect chance of re-infection, disease outcome. Survey of several other mutations showing reduction in antibody-mediated neutralization also revealed that similar disruption of H-bonding or salt-bridge or Van der Waals interaction might explain their phenotype. Analysis of GESS database indicated that N501Y, EK484 as well as these other mutations existed since March-April, 2020, might have evolved independently across the world and may keep accumulating, which could affect efficacy of vaccination and antibody-based therapies. Our analysis also indicated that these may spread in spite of current travel restrictions focused on few countries and evolve indigenously warranting intensification of surveillance for emerging mutations among all travellers as well as people in their dwelling zones. Meta-analysis of existing literature showed that repeat testing of travellers, contacts and others under scrutiny 7-11 days after the initial RT-PCR test may significantly help to contain the spread of emerging variants by catching false negative results. In addition, existing evidence calls for development of strain-specific tests, escalated sequencing and broadening the scope of surveillance including in hospitals and animal farms to contain the threat of emerging variants.

## Introduction

Along with loss of life, the socioeconomic impact of COVID-19 pandemic has been devastating for a large section of world population. Several assessments indicate that it would push a vast number of people into extreme poverty, hunger and derail progress towards improving of Human Development Indices.^1,2^ The spectrum of long-term health consequences of the SARS-COV-2 infection is quite a concern.^3,4^ In addition, it has significantly derailed immunisation programs ^5^ increasing the danger of resurgence of preventable infectious diseases. Amid all these, the emergence of the new SARS-COV-2 strains in Britain (VUI 202012/01 or B1.1.7) and South Africa (501Y.V2) is threatening to worsen situation.^6-9^ The UK variant was estimated to be 71% more infectious and may increase viral load as well. ^6, 7^ Emerging evidence also suggest that it may also lead to higher mortality. ^10^ It was detected in 31 countries outside UK by December, 30, 2020, itself. Almost at the same time, the variant from South Africa ^11^ spread very quickly in that country and had also been detected in travellers returning to UK. Another variant had also been also reported from Brazil in December, 2020. ^12^ While Japan reported detection of a novel variant from travellers returning from Amazonian states of Brazil in January, it was found to be different from the one circulating in Brazil. ^13^ However, understanding of the structural and molecular basis of improved fitness of these strains over parent Wuhan strain remains unclear.

As the number of cases with mutant strain kept piling up, many countries have stepped up testing and contact tracing of travellers and introduced fresh restrictions on mobility in December, 2020. However, it is quite likely that these variants actually emerged and transmitted much before that. For example, the B1.1.7 was traced back to samples collected on September 20, ^7, 14^ when none of these restrictions were in place. In fact, there has been considerable ambiguity about origin of these strains as well as the timeline of their arrival or evolution in different countries. Understanding these are very important to adopt appropriate strategy for containing the threat of these variants. In absence of adequate measures for surveillance, screening and testing, many of these strains are likely to transmit and cause havoc anywhere in the world. It is important to note that the leaky screening protocol for travellers based on body temperature and other symptoms failed us to arrest the spread of the pandemic. This kept happening in spite of emerging knowledge that many people can be asymptomatic and thus the aforementioned protocol had significant probability of false negative. For countries except China, COVID-19 was an imported disease, which could, in principle, be controlled with rigorous testing and isolation protocols at arrival. In fact, these were quite effectively implemented in countries such as Taiwan, Singapore, and New Zealand. A similar story of failure repeated with the D614G strain, which was predicted to be more infectious than the Wuhan strain, ^15,16^ and yet could spread swiftly across the world ^17^ since testing and isolation protocols were not revamped to detect the strain. It is important to note that variants with D796H, ΔH69/ΔV70 mutations detected in UK ^18^ as well as E484K and E417N mutations detected in Africa is suspected to have lower susceptibility towards highly neutralizing antibodies present in convalescent plasma ^19^. In addition to the highly infectious variants mentioned above, several other mutations have been detected in SARS-COV-2, which were found to be resistant to monoclonal antibodies ^20^ and, thus, may compromise the efficacy of vaccines and increase probability of re-infection. In fact, E484K mutation has already shown to be associated with re-infections in Brazil.^21^ Along with travel restrictions, the current strategy to contain cross-border transmission in most countries involves requirement of RT-PCR-negative status before boarding and generic RT-PCR test on arrival and sequencing of samples turning positive. In spite of these measures and even total travel ban in many cases, imported variants are still being detected in increasing numbers around the world. ^22,23^ Thus, in this study, we analyze the structural basis of most frequent mutations in the emerging variants as well as examine the literature to find best way to reduce false negative results for travellers and analyze the spatio-temporal trend of evolution SARS-COV-2 mutants of concern to recommend expanded retrospective surveillance through strain-specific testing and escalated sequencing.

## Method

### Structural implication of common mutations in emerging variants

The mutations found in three different variants of concern as per ECDC on its January 21, 2021 update ^24^ reported from UK (B1.1.7), South Africa (B1.351), Brazil and Japan (B1.1.28) ^25^ as well as two more strains related to infection clusters in Brazil ^12, 13^ were compared to identify most frequent mutations. Given that the most striking aspect of their phenotype has been increased infectivity and reduced susceptibility to antibodies, we chose to analyze the effect of two most common mutations identified in the spike protein on interaction with the human ACE2 receptor and a neutralizing antibody. The high resolution crystal structure of the complex of the receptor binding domain (RBD) of the spike protein in complex with the human ACE2 receptor (hACE2) (PDB ID: 6M0J, resolution 2.6 Å) ^26^ and that with the neutralizing antibody P2B-2F6 (PDB ID: 7BWJ, resolution 2.65 Å) ^27^ were used for these analysis. Effect of individual mutations were examined analyzed by estimating the difference of biding free energies (ΔΔG) using MutaBind2 server (https://lilab.jysw.suda.edu.cn/research/mutabind2/). ^28^ The PDB file of the mutant complex generated by for the analysis was further used to analyze any significant deviation in the dihedral angles in the Ramachandran Plot using Ramachandran Plot Server (https://zlab.umassmed.edu/bu/rama/) ^29^ including glycine and proline residues. These structures were analyzed using RASMOL^30^ (version 2.7.5.2) to examine the effect of mutations on intra- and inter-molecular interactions.

### Spatio-temporal evolution of SARS-COV-2 mutations

The SARS-COV-2 variant database GESS ^31^ was searched to find temporal trend of deposition and geographical distribution of single nucleotide variants (SNV) including those coding for E484 and N501Y ^32^ spike protein mutations as well as co-occurring SNVs that have been identified in emerging variants found in UK, South Africa, Brazil and Brazil-related variant outside Brazil (Japan). In addition, SNVs of spike protein RBD residues involved in H-bonding or salt bridge (K417, N440, K444, N448, N487, Q493) as well as significant Van der Waals interactions (L452, F453, A475, V483, F486, F490) with residues of neutralizing antibodies as revealed by high resolution crystal structures ^27, 33^ were also included for analysis. Moreover, along with ‘escape mutations’ in the spike protein RBD, others like D796H, H655Y that have been shown to be markedly resistant to antibodies ^18, 34-37^ were examined for their spatio-temporal evolution.

### Meta-analysis of rate of false negative results in RT-PCR test for SARS-COV-2

A search on the Pubmed with the keywords ‘SARS-COV-2 + RT-PCR + false negative’ was performed to find published and preprint articles. In addition, articles including preprints cited in earlier literature surveys ^38-40^ on false negative RT-PCR rates were also included. These were screened to select only original articles reporting results from at least three independent samples. These articles were manually examined to find respective screening modalities (population-based or symptom-based or upon arrival at hospitalization), types of samples, target genes and protocols followed, percentages of false negatives in initial RT-PCR along with the basis of calling false negatives as well as interval between initial RT-PCR and reassessment to identify false negatives. Correlation between RTPCR false negative rates and the interval between initial test and reassessment through repeat RTPCR or CT scan were examined using Pearson correlation using GraphPad Prism 7. The screening of a traveller at arrival is similar in nature with population-based screening since in both cases a subject may already be infected for a while and have had a false negative result before boarding or may have been inoculated during the boarding and air travel or may still be uninfected. The concern in both cases is the ability to catch very recent and asymptomatic infections with low viral load. It is safe to assume that barring sampling and sample processing issues or RT-PCR protocol itself, most people returning false negative results may have low viral load such as those with very recent infections. The median interval between inoculation and symptom onset has been shown to be 5.1 days (95% CI, 4.5 to 5.8 days), ^41^ which was also used by Kucirka *et al* ^39^ for analysis variation of false negative results with sampling time. So, the median reassessment interval for studies reporting symptom-based testing were adjusted by an additional five days to compare them with false negatives in population-based or traveller screening associated with very recent infection. The false negative RT-PCR rates of symptom-based testing was plotted against adjusted interval together with that of population-based screening to generate the expected variation of false negative rate in the initial RT-PCR test with respect to time interval to reassessment and find the best strategy to catch these false negative results.

## Results

### Identification of most frequent mutations in emerging variants

Table 1 summarizes changes in amino sequences identified in five different emerging variants of SARS-COV-2. While no mutation was found to be present in all of them, N501Y and E484K mutants were found in four out of five variants. So, these two were chosen to analyze their plausible consequences on interaction with the hACE2 receptor and the neutralizing antibody P2B-2F6.

**Table 1:**
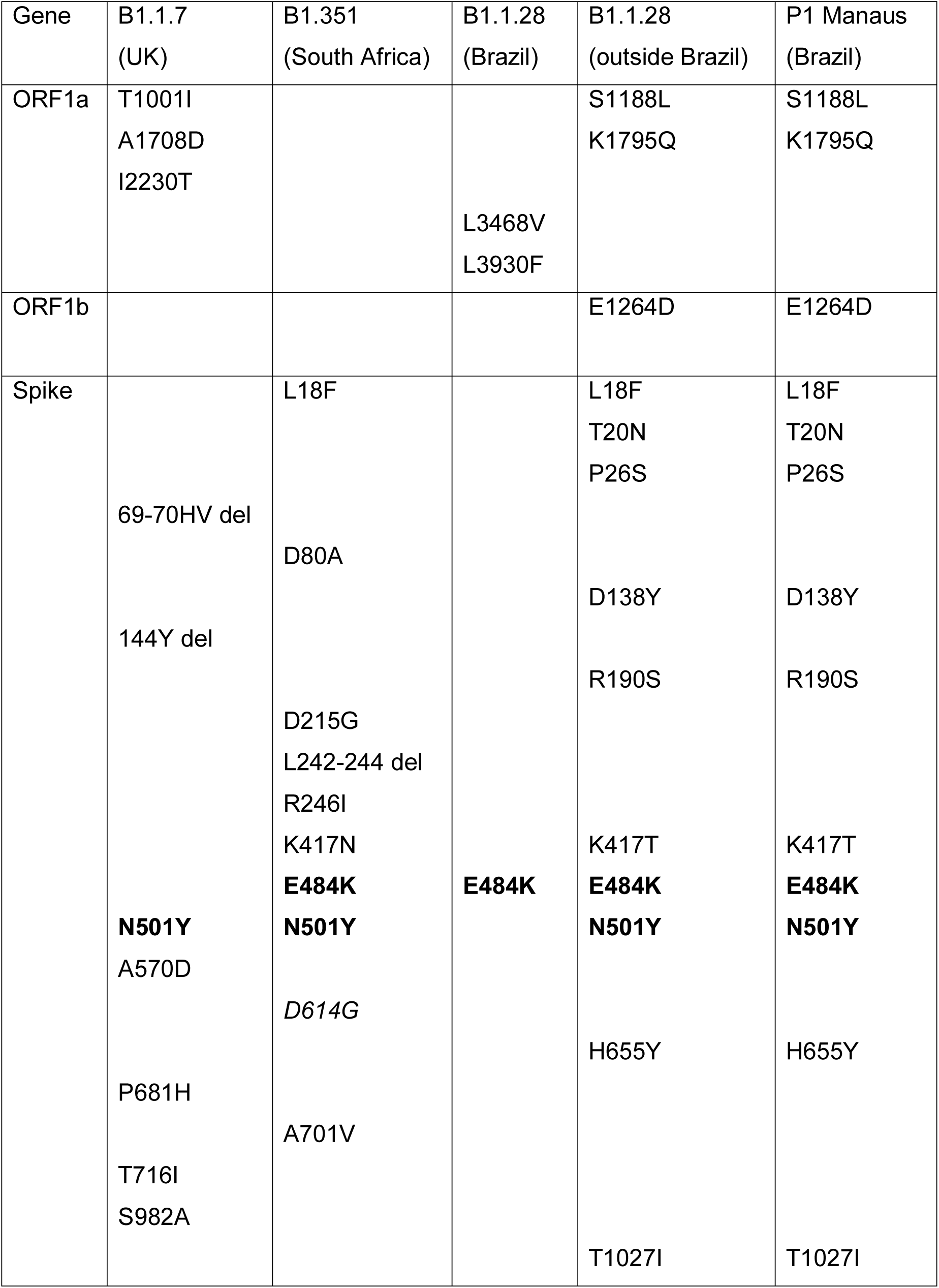
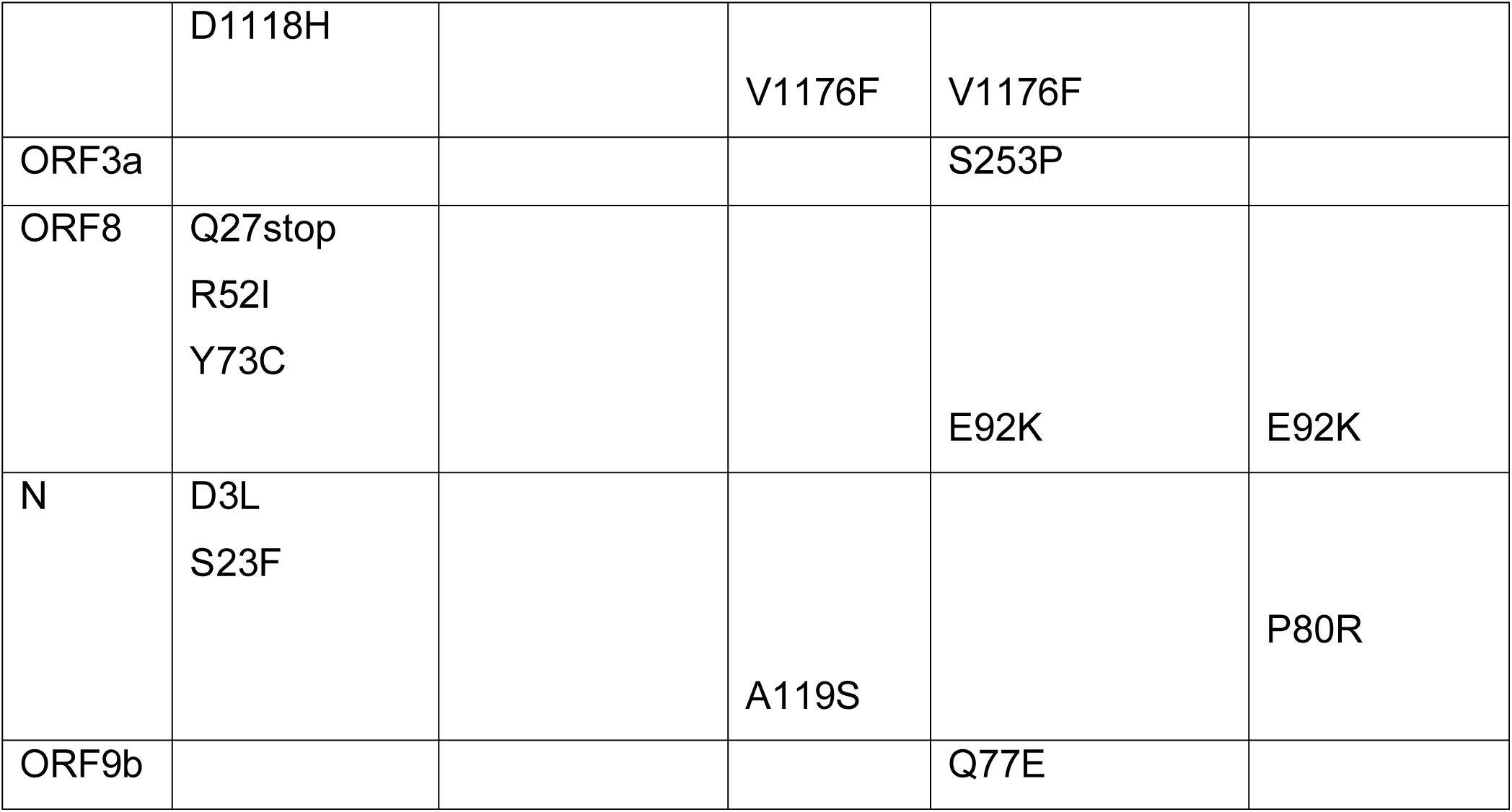
Comparison of non-synonymous mutations identified in emerging SARS-COV-2 strains.

### Effect of N501Y and E484 mutations on interaction with hACE2 receptor

The *in silico* analysis through MutaBind2 algorithm estimated ΔΔG values of 0.18 and 0.61 kcal/mole, respectively, for hACE2 complexes of E484K and N501Y mutants. Both mutations were predicted to be non-deleterious for the binding interaction. All PDB files generated for the point mutation can be found in supplementary material. The analysis of dihedral angles showed that the original crystal structure (PDB: 6M0J) had 765 residues in highly preferred conformation, 13 residues in preferred and 7 residues in questionable conformation according to the Ramachandran plot (Figure S1A). Residues with questionable conformation included three residues from the spike protein RBD and four residues from the ACE-2 receptor. Similar analysis of showed 767, 15 and 5 residues respectively, in highly preferred preferred and questionable conformation in the N501Y mutant complex (Figure S1B). The E484K mutant complex also had 767, 15 and 5 residues respectively, in highly preferred, preferred and questionable conformation (Figure S1C). In case of mutants, two residues in questionable conformation were from the ACE-2 receptor and three residues from the spike protein RBD. All residues in questionable conformation were glycine residues. Analysis showed no significant change in secondary structure of the spike protein RBD upon mutation (data not shown).

The analysis of residues involved in the interaction between hACE2 receptor and the spike protein RBD showed that N501 residue is present in the binding surface. As was shown earlier, ^26^ it was involved in interaction with the Tyr41, Lys353, Gly354 and Asp355 of hACE2 receptor. It was noted that the distance between the Y41 side chain oxygen and the side chain amide nitrogen of N501 was 3.43Å (Figure 1A), indicating a weak H-bond. Residues around this region from the RBD and the hACE2 receptor were involved in several other H-bonds that are mentioned in Table 2 and shown in Figure 1A. The Y501 residue in the N501Y mutant, on the other hand, had no H-bonding interaction with the Y41 residue (Figure 1B). But it seemed to form a strong H-bond (2.79Å) with the side chain amine of the K353 residue of the hACE2 receptor. The K353 residue was involved in a relatively weak intra-molecular H-bond (3.17Å) with one of the side chain oxygen of the D38 residue in the complex with the Wuhan strain RBD. However, this distance reduced to 2.5Å indicating a strengthening of H-bond in the N501Y mutant. The steric bulk of the 501Y side chain was also found to cause a flip in the Q498 side chain in such a manner that the distance between the Q498 side chain amide nitrogen and the Q42 side chain amide oxygen reduced from 3.38Å for the Wuhan strain to 3.06Å in the N501Y mutant. In addition, while the distance between the backbone carbonyl oxygen of G496 residue and the N501 side chain nitrogen was 3.4Å, the 501Y side chain oxygen was placed just 2.5Å from the G496 carbonyl in the N501Y mutant.

**Table 2:** Inter and intra-molecular H-bond distances in the human ACE2 receptor in complex with spike protein RBD variants.

| Interacting atoms <sup>a</sup> | N501 <sup>b</sup><br>Distance (Å) | N501Y <sup>c</sup><br>Distance (Å) |
| --- | --- | --- |
| (hACE2) Y41OH-N501OD1(RBD) | 3.43 | - |
| (hACE2) K353-Y501OH (RBD) | - | 2.79 |
| (hACE2) Q42OE1-Q498NE2 (RBD) | 3.38 | 3.06 |
| (hACE2) Q42NE2-Y449OH (RBD) | 2.79 | 2.8 |
| (hACE2) D38OD2-Y449OH (RBD) | 2.69 | 3.01 |
| Q496O (O)-N501ND2 (RBD)<sup>a</sup> | 3.4 | - |
| Q496O (O)-Y501OH (RBD) | - | 2.5 |
| (hACE2)D38OD1-K353NZ (hACE2) | 3.17 | 2.5 |

<sup>a</sup> Atoms in bracket represent peptide backbone amide oxygen or nitrogen.
<sup>b</sup> Based on PDB ID: 6M0J (Lan J, et al. 2020)
<sup>c</sup> Based on computationally generated structure through MutaBind2 server in this work.

**Figure 1.**
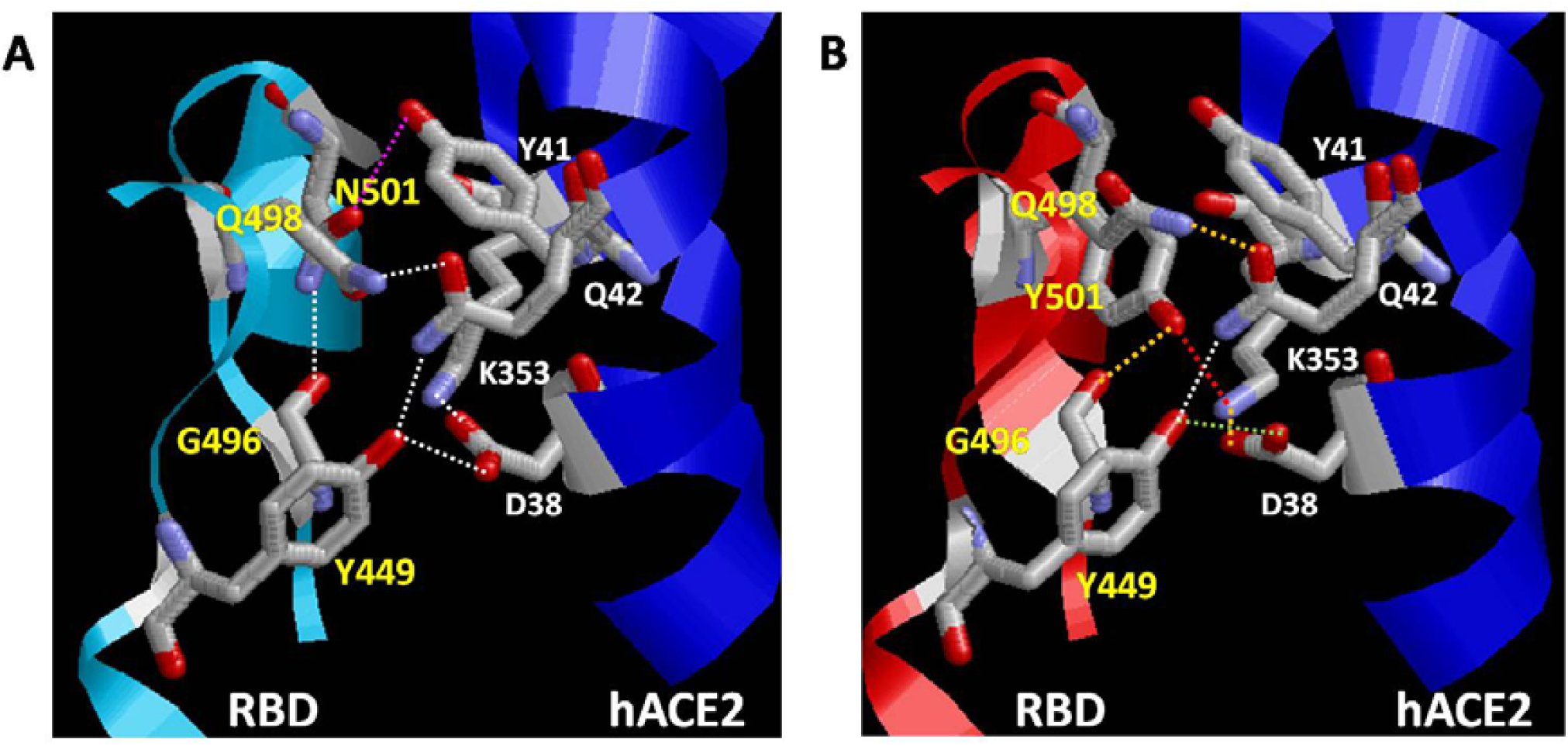
Intra-and intermolecular H-bonding in the (A) Wuhan strain and (B) N501Y mutant of SARS-COV-2 spike protein receptor binding domain in complex with human ACE2 receptor. Blue ribbon indicates human ACE2 protein chain while cyan ribbon in panel A and red in panel B indicate spike protein RBDs of Wuhan strain and N501Y mutants, respectively. Red, pink, orange and green dotted lines indicate new, lost, shortened and lengthened H-bond, respectively. Inter-atomic distances indicated here are mentioned in Table 2.

The E484 residue is located at the surface of the spike protein RBD (Figure S2A) and was not found to be involved in any Van der Waals, H-bonding or salt-bridge interaction with the hACE2 receptor. The E484K mutation resulted in reversal of the charge of the surface exposed residue. In addition, the charged amine group of the lysine side chain was also found to be positioned on top of the aromatic ring of the F490 residue (Figure S2B). This could result in pi-cation interaction and may affect any intermolecular interaction that involves these two residues.

### Effect of N501Y and E484 mutations on interaction with neutralizing antibody

The MutaBind2 analysis estimated a ΔΔG of 1.74 kcal/mole to be associated with the E484K mutation of the spike protein for complex formation with the P2B-2F6 neutralizing antibody. The mutation was predicted to be deleterious for the binding with P2B-2F6. The analysis of dihedral angles showed that the original crystal structure (PDB: 7BWJ) had 598 residues in highly preferred conformation, 25 residues in preferred and 7 residues were in questionable conformation according to the Ramachandran plot (Figure S3A). Residues with questionable conformation included 2, 3 and 2 residues, respectively, from the spike protein RBD, heavy and light chains of the antibody. Similar analysis showed 597, 26 and 7 residues respectively, in highly preferred, preferred and questionable conformation for the E484K mutant complex (Figure S3B). The N501Y mutant complex also had 600, 23 and 8 residues respectively, in highly preferred, preferred and questionable conformation (Figure S3C) with 2, 4 and 2 residues in questionable conformation belonging to spike protein RBD, heavy and light chains of the antibody, respectively. All residues in questionable conformation were glycine residues. Analysis showed no significant change in secondary structure of the spike protein RBD upon mutation (data not shown).

The E484 residue is involved in an H-bond interaction with the Y34 side chain OH of the heavy chain as well as a salt-bridge interaction with the R112 side chain of the light chain of the antibody (Figure 2A). Inter-atomic distances associated with intermolecular H-bond, salt-bridge and Van der Waals interactions in the vicinity of the E484 residue are mentioned in Table 3. As shown in Figure 2B, the E484K mutation flips the positively charged K484 residue away from the positively charged R112 residue of the heavy chain. Along with loss of the salt-bridge interaction, this also removes the H-bond involving the Y34 side chain of the light chain. The K484 side chain instead forms an intra-molecular H-bond (2.74Å) with the backbone carbonyl oxygen of the G485 residue. Analysis of the interface between the RBD and antibody in the vicinity of the 484 residue revealed that the mutation also weakens the H-bond (2.5Å to 3.02Å) between the 484 backbone carbonyl oxygen and side chain amide nitrogen of N33 residue of the light chain of the antibody (Figure 2B). In addition, the H-bonds involving Y27 (heavy chain) and G447 (RBD) as well as H54 (heavy chain) and N450 (RBD) were found to lengthen whereas that involving S31 (heavy chain) and Y449 (RBD) was found to shorten. The Van der Waals interaction involving the F490 residue (RBD) and V105-V106 of the heavy chain was found to be relatively unaffected.

**Table 3:** Inter-atomic distances at the interface of complex between P2B-2F6 neutralizing antibody and spike protein RBD variants

| Interacting atoms <sup>a</sup> | Nature of interaction | E484 Distance (Å) <sup>b</sup> | E484K Distance (Å) <sup>c</sup> |
| --- | --- | --- | --- |
| (nAb-L) N33ND2-E484(O) (RBD)<sup>b</sup> | H-bond | 2.5 | 3.02 |
| (nAb-L) N33ND2-K484(O) (RBD) | H-bond | - |  |
| (nAb-L) Y34OH-E484OE2 (RBD) | H-bond | 2.47 | - |
| (nAb-L) Y34OH-K484NZ (RBD) |  | - | 7.07 |
| (RBD) G485(N)- K484NZ (RBD) | H-bond | - | 2.74 |
| (nAb-H) R112NH1-E484OE2 (RBD) | Salt bridge | 3.57 | - |
| (nAb-H) R112NH1-E484OE1 (RBD) | Salt bridge | 3.83 | - |
| (nAb-H) Y27OH-G447(O) (RBD) | H-bond | 2.93 | 3.21 |
| (nAb-H) H54NE2-N450OD1 (RBD) | H-bond | 2.83 | 3.01 |
| (nAb-H) S30(O)-N450ND2 (RBD) | H-bond | 3.28 | 3.28 |
| (nAb-H) S31(O)-Y449(N) (RBD) | H-bond | 3.11 | 2.88 |
| (nAb-H) V106CG1-F490CE2 (RBD) | Van der waals | 4.38 | 4.06 |
| (nAb-H) V105CG2-F490CZ (RBD) | Van der waals | 3.79 | 3.85 |
| (nAb-H) V106CA-F490CZ (RBD) | Van der waals | 4.04 | 4.14 |
| (nAb-H) V105C-F490CZ (RBD) | Van der waals | 4.05 | 4.02 |

<sup>a</sup> Light and heavy chains of the neutralizing antibody P2B-2F6 indicated by nAb-L and nAb-H, respectively.
<sup>b</sup> Based on PDB ID: 6M0J (Ju B, et al. 2020)
<sup>c</sup> Based on computationally generated structure through MutaBind2 server in this work.
<sup>d</sup> Atoms in bracket represent peptide backbone amide oxygen or nitrogen

**Figure 2.**
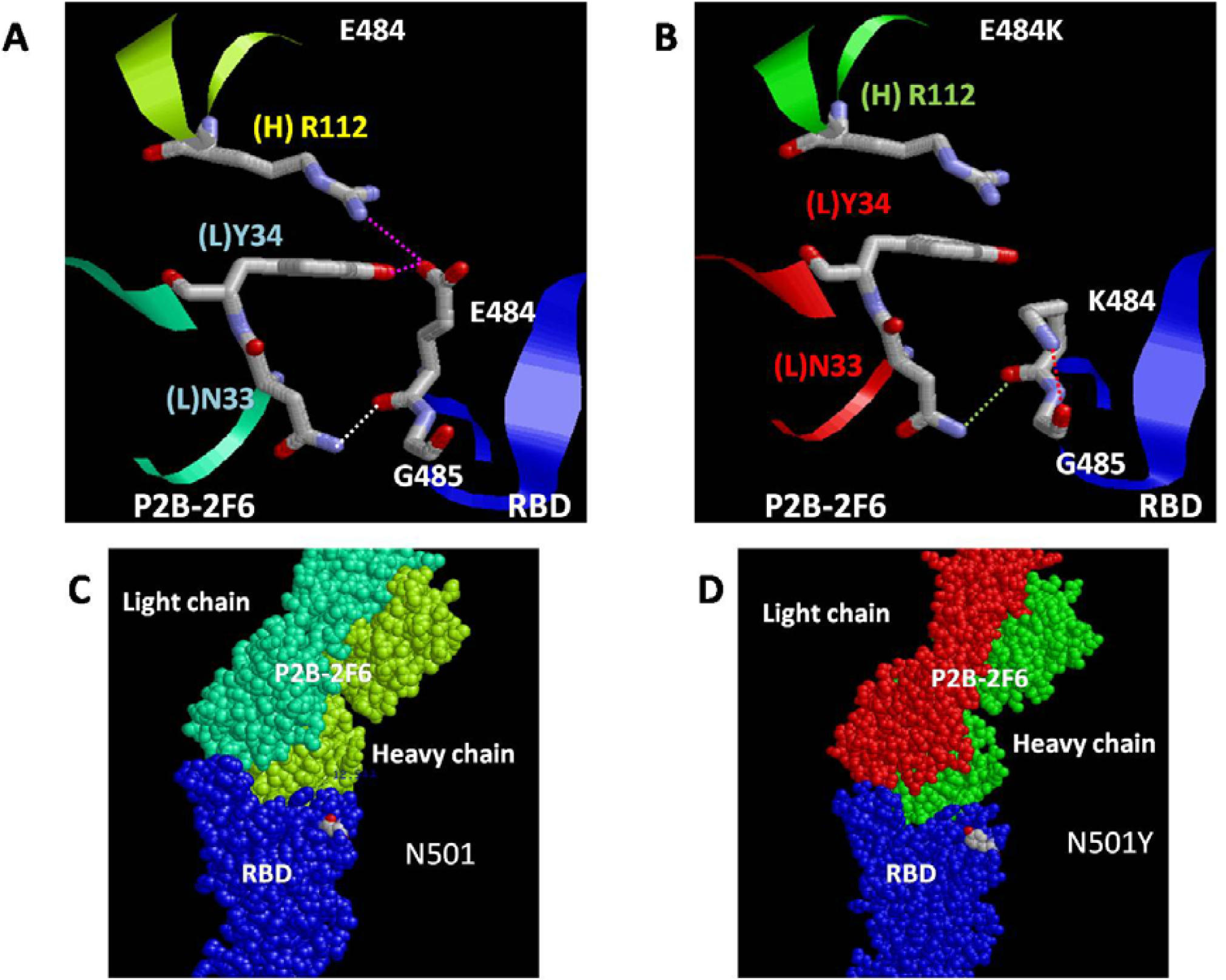
Intra-and intermolecular H-bonding involving the (A) Wuhan strain and (B) E484K mutant of SARS-COV-2 spike protein receptor binding domain in complex with P2B-2F6 neutralizing antibody. Pink, red and green dotted lined indicate lost, new and weakened H-bonds, respectively. Blue, yellow and cyan ribbons indicate spike protein RBD, heavy chain (H) and light chains (L) of the P2B-2F6 neutralizing antibody in the Wuhan strain complex. Heavy and light chains in the E484K complex are represented by green and red ribbons. Spacefill models showing positions of (C) N501 and (D) Y501 residues of spike protein RBD in respective complexes with P2B-2F6 neutralizing antibody. The colour scheme for RBD and light chain, heavy chain of the antibody is same as that in A and B.

### Spatio-temporal evolution of SARS-COV-2

The list of different SNVs searched in the GESS database along with their monthly as well as country-wise absolute numbers reported so far can be found in Supplementary info. As has been noted earlier, the frequency of detection of D614G mutation increased sharply from 21.74% in February to 70.53% in March, crossed 99% October, 2020 and ever since remained almost 100% across the world till date (Figure 3A). It was noted that both N501Y and E484K mutations were first reported in March, 2020 (Figure 3B). While their percentage increased relatively slowly, a spurt was noted since September, 2020. Interestingly, L18F mutation that was found in both South Africa (B1.351) and Brazil variants (P1 Manaus), could be traced back to February, 2020 and showed a sharp increase, as seen for D614G, after July, 2020. L18F mutation was significantly more abundant (> 30%) than N501Y and E484K mutations (<1%) in December, 2020. Other spike protein SNVs that have either been found in emerging variants (Table 1) or shown to be less susceptible to neutralizing antibodies were also found to be reported in the GESS database since long (Figure 3C and 3D). In fact, except R190S, S982A all these mutations were found to be reported back in April, 2020 or earlier.

**Figure 3.**
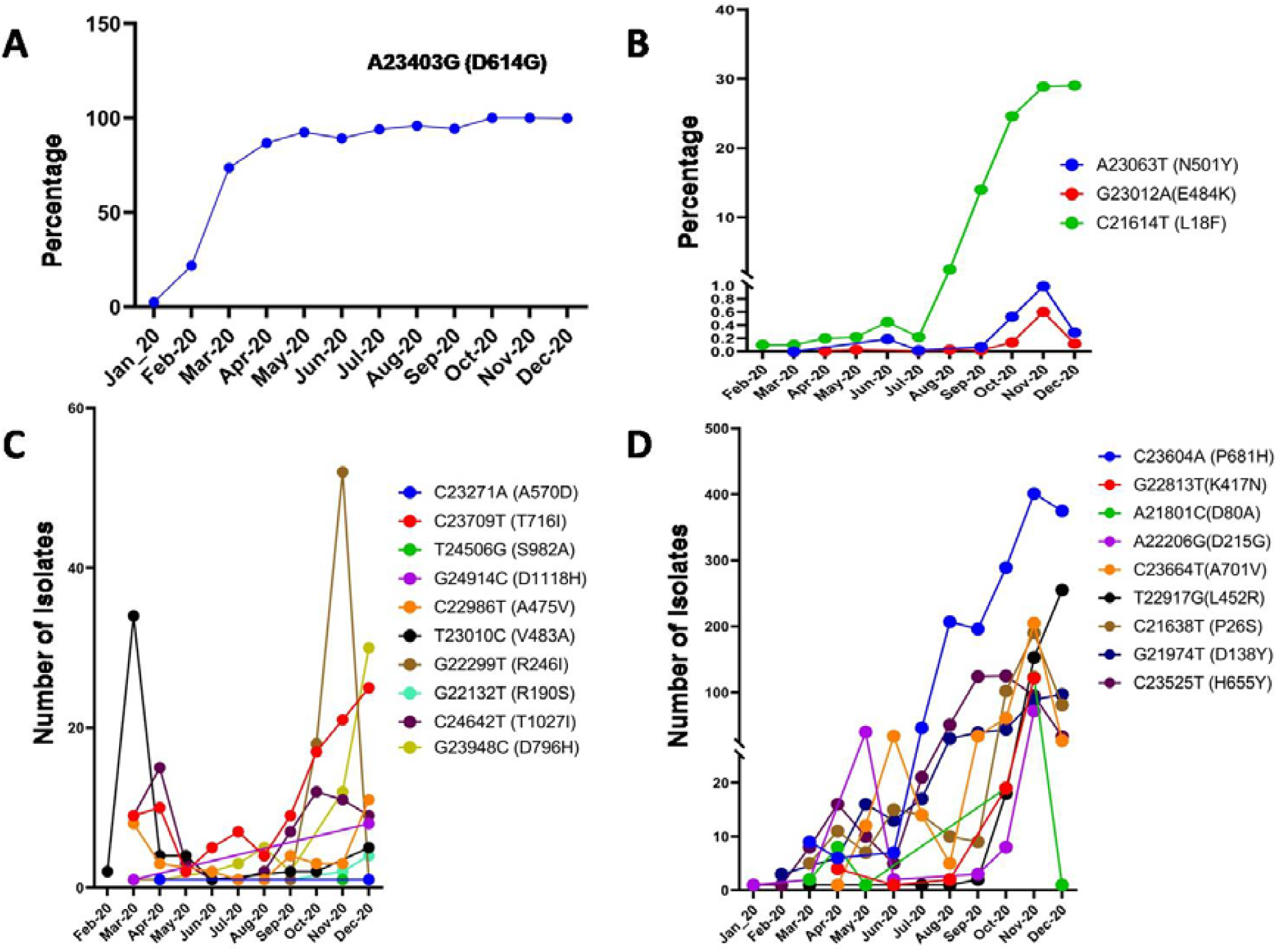
Monthly variation in percentage of SNVs representing (A) D614G and (B) N501Y, E484K and L18F mutations in spike protein in GESS database. Monthly variation in number of sequences with SNVs representing (C) R190S, R246I, A475V, V483A, A570D, D716I, S982A, T1027I, D1118H and (D) P26S, D80A, D138Y, D215G, D417N, H655Y, P681H, A701V in GESS database.

Analysis of co-occurring SNVs show that while both N501Y and E484K mutation has been reported since long, their co-occurrence (N501Y-E484K) in GESS database could only be identified since October, 2020. Nevertheless, as the sequence of events show (Figure 4C), this precedes the first reported sequencing of B1.351 (South Africa), B1.1.28 (outside Brazil) and P1 Manaus (Brazil) variants as well as imposition of travel restrictions from UK, all of which happened December 2020 onwards. It was interesting to note that even according to the latest GESS database, the co-occurrences of N501Y and E484K was found only in samples from South Africa so far (see Supplementary info) while Brazilian variants harbouring them has already been sequenced. This indicated that absence in GESS should not be taken as any evidence for absence of a mutation or variant in a particular country or time. The analysis of other SNVs on RBD residues (440, 444, 448, 452, 453, 475, 483, 484, 486, 487, 490 and 493) as well as other spike protein residues (R346, H655Y) implicated in evolution of mutations with reduced susceptibility to neutralizing antibodies showed (Figure S4 and Figure 3D) that several mutations appeared throughout the pandemic and well before December, 2020.

**Figure 4.**
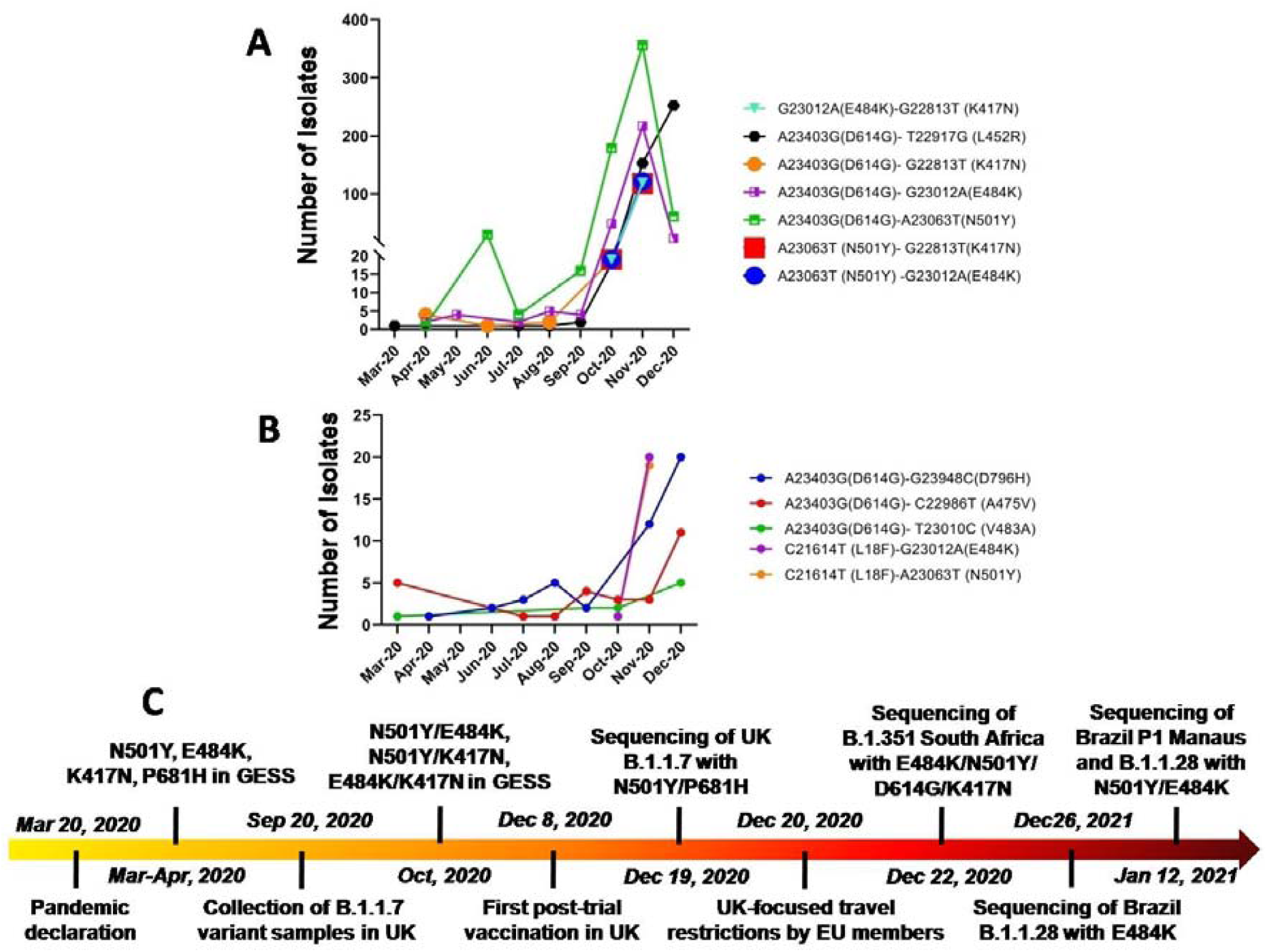
Monthly variation in number of sequences with co-occurring SNV pairs involving (A) E484K or N501 or D614 mutations and (B) L18F and D614G mutation as reported in GESS database. (C) Sequence of events related to emergence mutations of concern.

The cumulative country-wise distribution of SNVs (Figure 5) showed that while N501Y mutation was detected in 8 countries, N501T, N501I and N501H mutations were detected in 12, 2 and 1 countries, respectively. E484K mutation was reported in 18 countries, whereas E484Q and E484A were reported in 11 and 4 countries, respectively. P681 mutation that co-occurs with N501Y in the B1.1.7 UK variant was present in 27 countries (Figure 5S). Mutations associated with reduced susceptibility to neutralizing antibodies and escape such as K417N, H655Y, N440K, D796H, N450K were reported in 7, 23, 11, 5 and 4 countries, respectively. L452R and L452M were detected in 22 and 6 countries respectively. Other mutations associated with significant changes in ability to participate in H-bond or salt-bridge or Van der Waals interactions, such as K444R, K444N, N487D, R346I, R346S, R346K, Y453F, F490L, F490S, V483F and D138Y were detected in 3, 4, 1, 3, 1, 12, 5, 2, 7, 11 and 22 countries, respectively (Figure S6). While most of these mutations were reported from Europe, particularly, Great Britain area, South Africa, Brazil and USA, several other countries in different parts of the world showed presence of multiple mutations.

**Figure 5.**
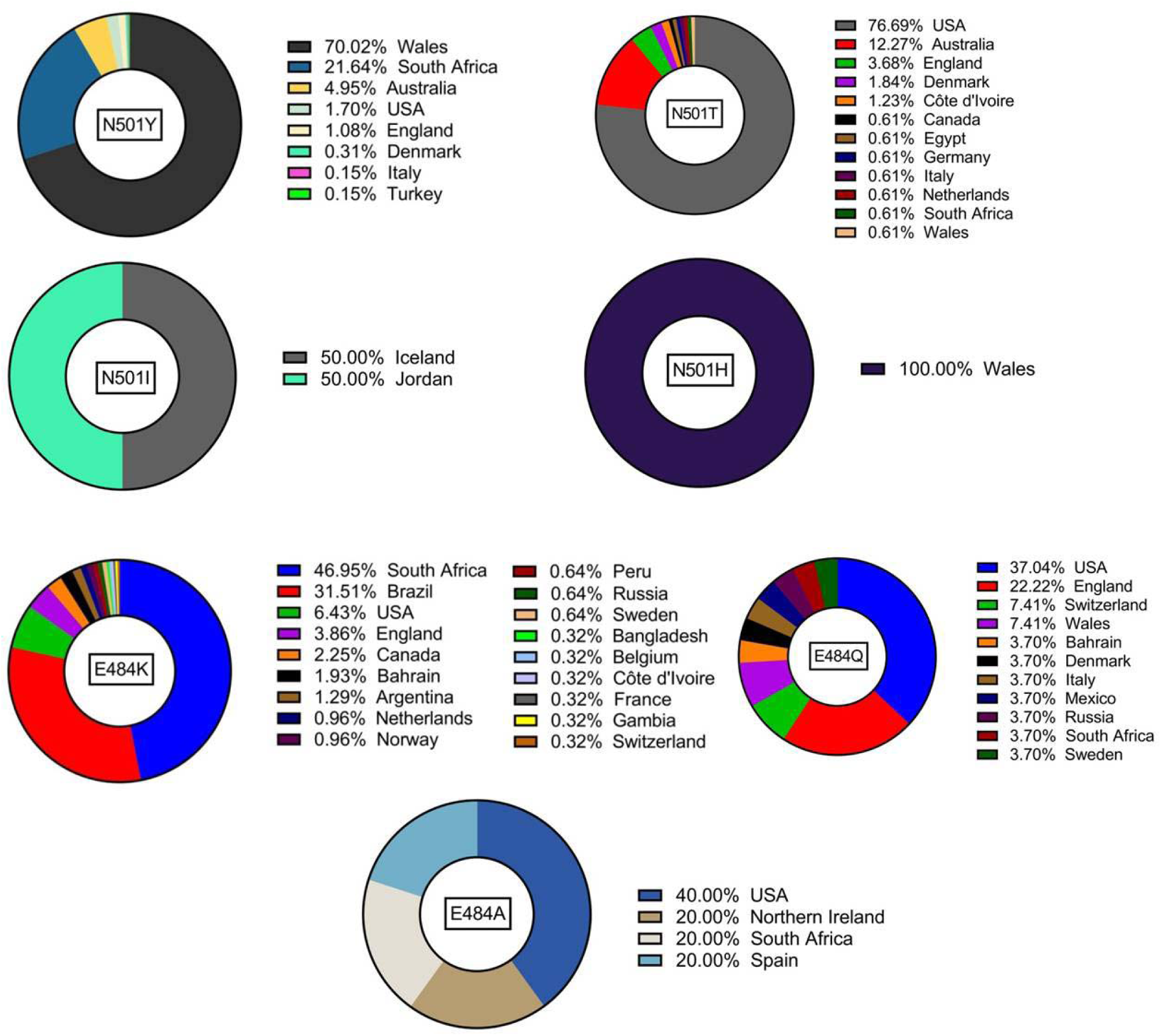
Country-wise relative abundance of N501Y, N501T, N501Y, N501H and E484K, E484Q and E484A mutations as reported in GESS database.

### Factors affecting false negative results in RTPCR test for SARS-COV-2

A search on the Pubmed with the keywords ‘SARS-COV-2 + RT-PCR + false negative’ returned 223 results including published and preprint articles. Along with original articles found in them, published articles and preprints cited in review articles in Pubmed were also included. These articles were screened to identify articles reporting analysis of at least three samples clearly mentioning subject selection criteria for initial RT-PCR, basis for calling false negative and percentage of false negative results. This yielded 24 original articles including four on population-based screening, ^42-45^ ten on symptomatic test ^46-55^ and ten on tests performed at hospitalization ^56-65^ as summarized in Tables 4, 5 and 6 respectively. The percentage of false negative results for the initial RT-PCR test that were reported in these studies ranged between 2-100%. The study reporting 100% false negative result was performed on day one at hospitalization. The weighted average of false negative rates for the initial RT-PCR analysis in population screening (122399 samples), symptomatic testing (1692 samples) and hospital-based testing (2742 samples) were estimated to be 8.41%, 9.97% and 17.45%, respectively. The overall false negative rates combining all of them were estimated to be 8.58%. Among population-based sampling studies, it was noted that as the time gap between initial RT-PCR and the repeat RT-PCR or CT scan to confirm results increased from 1 day to 4 days to 6.1 days on average, the detection rates of false negatives increased linearly (Figure 6A, R^2^ = 0.878) from 2% to 4% to 9.3% (Table 4). ^42-44^ Data from the population-based screening study with sero-positivity-based false negative calling ^45^ could not be used in analysis since a significant number of people may not become sero-positive. On the other hand, false negative rate for symptom-based RT-PCR tests decreased linearly (Figure 6B, R^2^ = 0.674) with re-assessment interval wherever they were mentioned (see Table 5). ^46, 48, 51, 54, 55^ While three studies on hospital-based testing mentioning the interval to reassessment ^57, 58, 63^ showed no such linearity between reassessment interval and false negative rate (Figure 6C, Table 6), repeat RT-PCR testing of nasal swabs after a median interval of 7.5 days reported detection of highest fraction of initial false negative RT-PCR results. ^58^ Plot of RTPCR false negative detection on reassessment against adjusted interval between initial RT-PCR and reassessment combining population-based screening ^42-44^ and symptom-based testing studies ^46, 48, 51, 54, 55^ showed a peak between 7-8 days. It should be noted that although desirable, the interval between symptom onset and initial RT-

**Table 4:** False negative rates for SARS-COV-2 RT-PCR test reported for population-based screening

| Sample number<sup>a</sup> | Sample type | RT-PCR targets/kits | False negative (%) | Basis of false negative calling |
| --- | --- | --- | --- | --- |
| 100001<br>(Kanji JN et al, 2021) | Nasopharyngeal<br>Oropharyngeal<br>Deep nasal-turbinate swabs<br>Endotracheal-aspirates<br>Bronchoalveolar lavages | Laboratory designed testing Kit.<br>Target genes: E and RdRp | 9.3% | Two repeat RT-PCR test with target genes (E, N1 and N2).<br>Mean time between two RT-PCR tests is 6.1 days. |
| 20912<br>(Long DR, et al 2020) | Nasopharyngeal | RT-PCR was performed targeting the N, RdRp, S, and E and ORF1ab | 4% | Repeat RT-PCR test withing 7 days.<br>Mean interval between last negative and then a positive test is 4.0 days (SD-2.0 days). |
| 1014<br>(Ai T, et al, 2020) | Throat swabs | TaqMan one step RT-PCR kits from shanghai huirui biotechnology Co Ltd,<br>Target genes are orf1ab, N and E genes. | 2% | Reconfirmed positive on basis of Chest CT report.<br>Median time between chest CT and RT-PCR was 1 days (0-7 days range) |
| 2021<br>(Baron R C, et al, 2020) | Nasopharyngeal and oropharyngeal swabs | Not mentioned | 13% | Seropositivity |

<sup>a</sup> References mentioned in bracket.

**Table 5:**
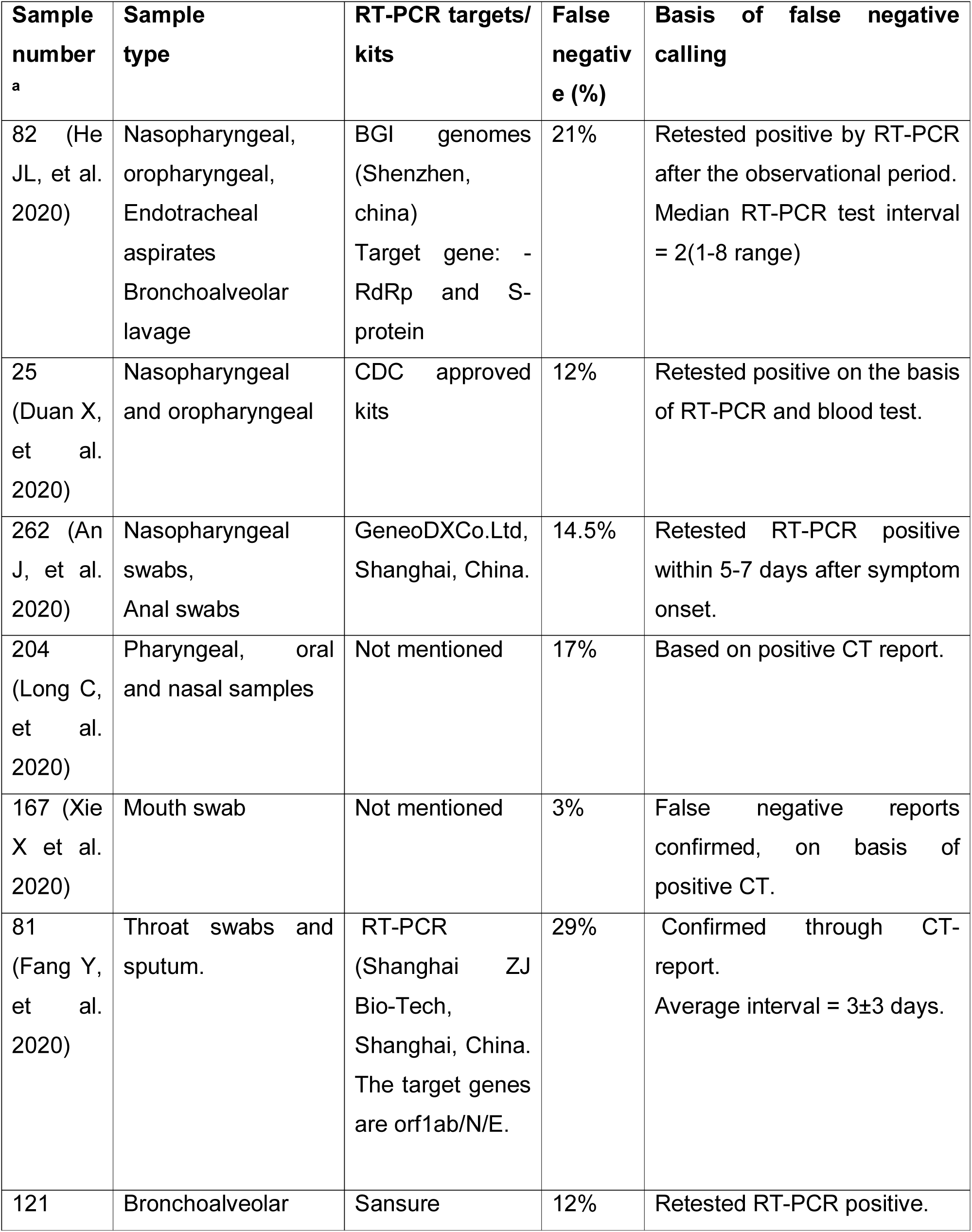
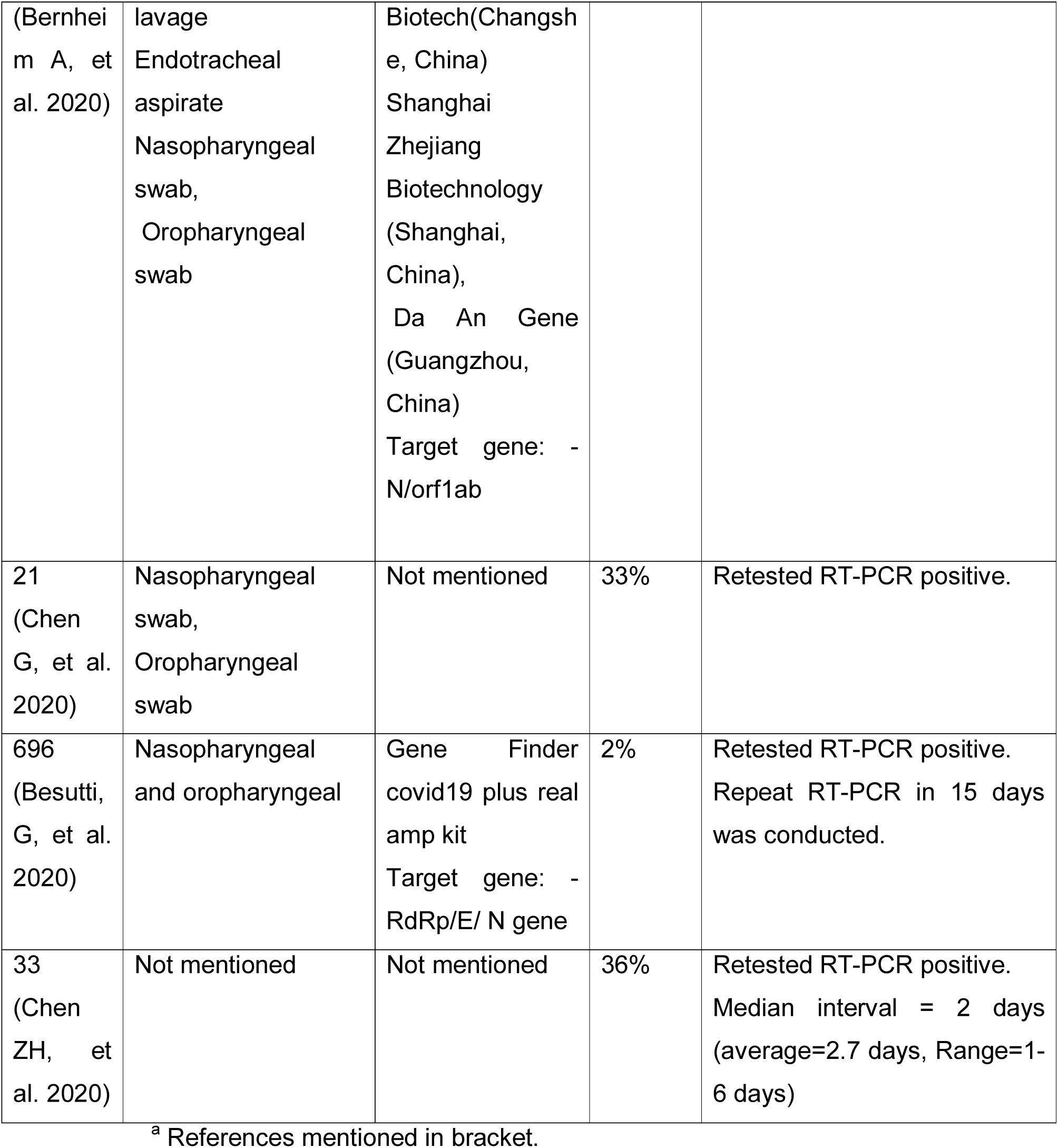
False negative rates for SARS-COV-2 RT-PCR test reported for symptom-based testing

**Table 6:**
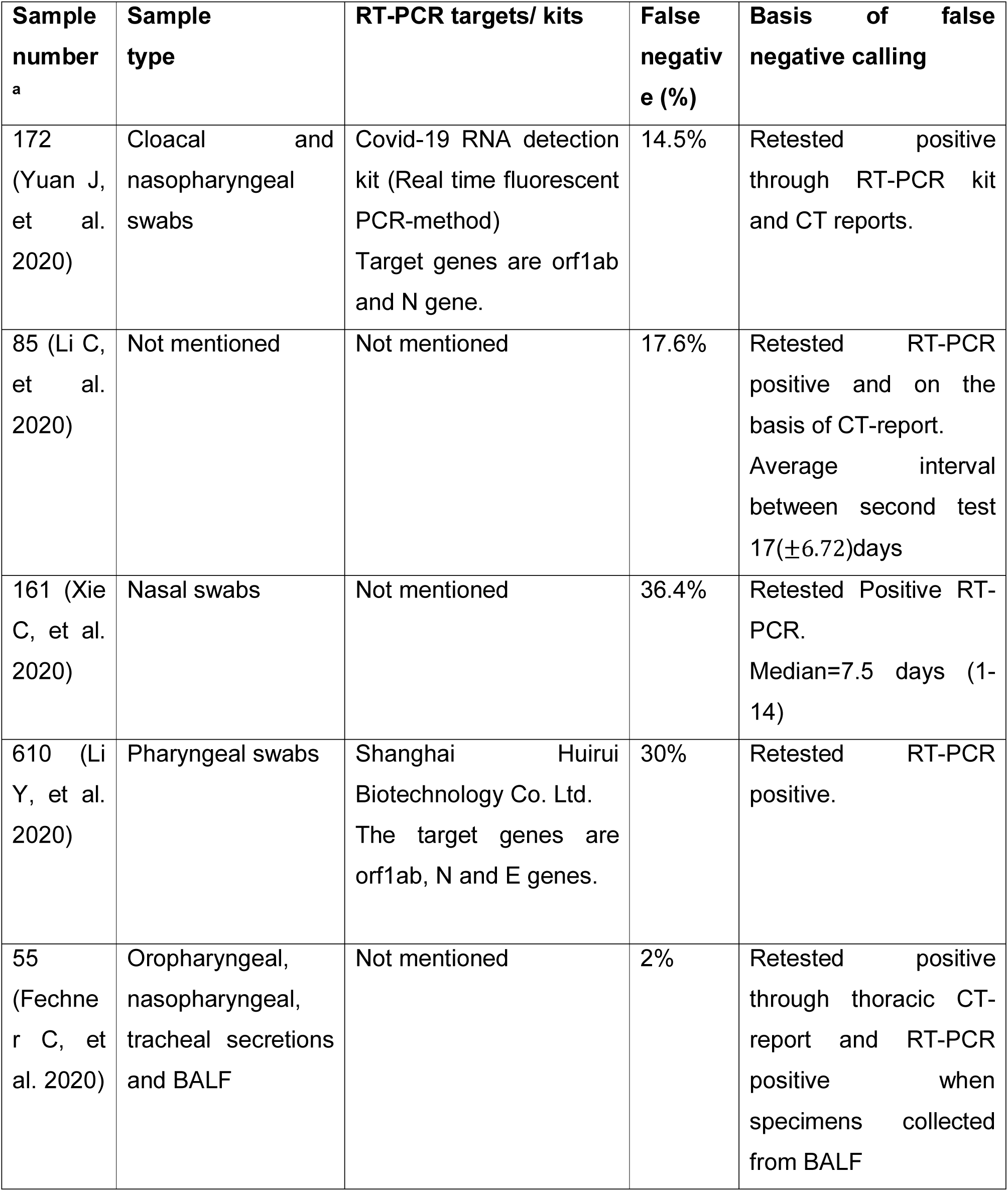
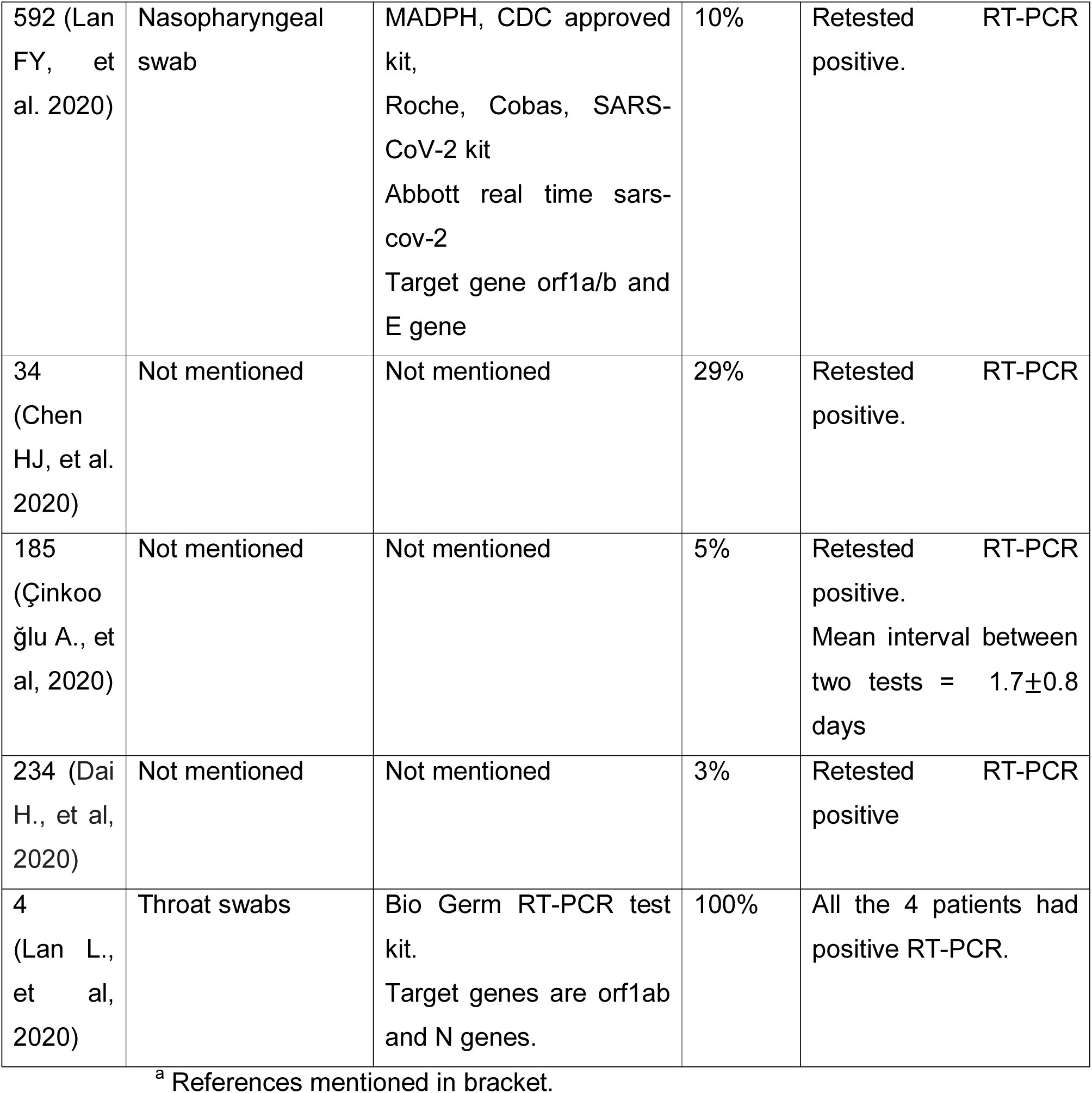
False negative rates for SARS-COV-2 RT-PCR test reported for hospital-based

**Figure 6.**
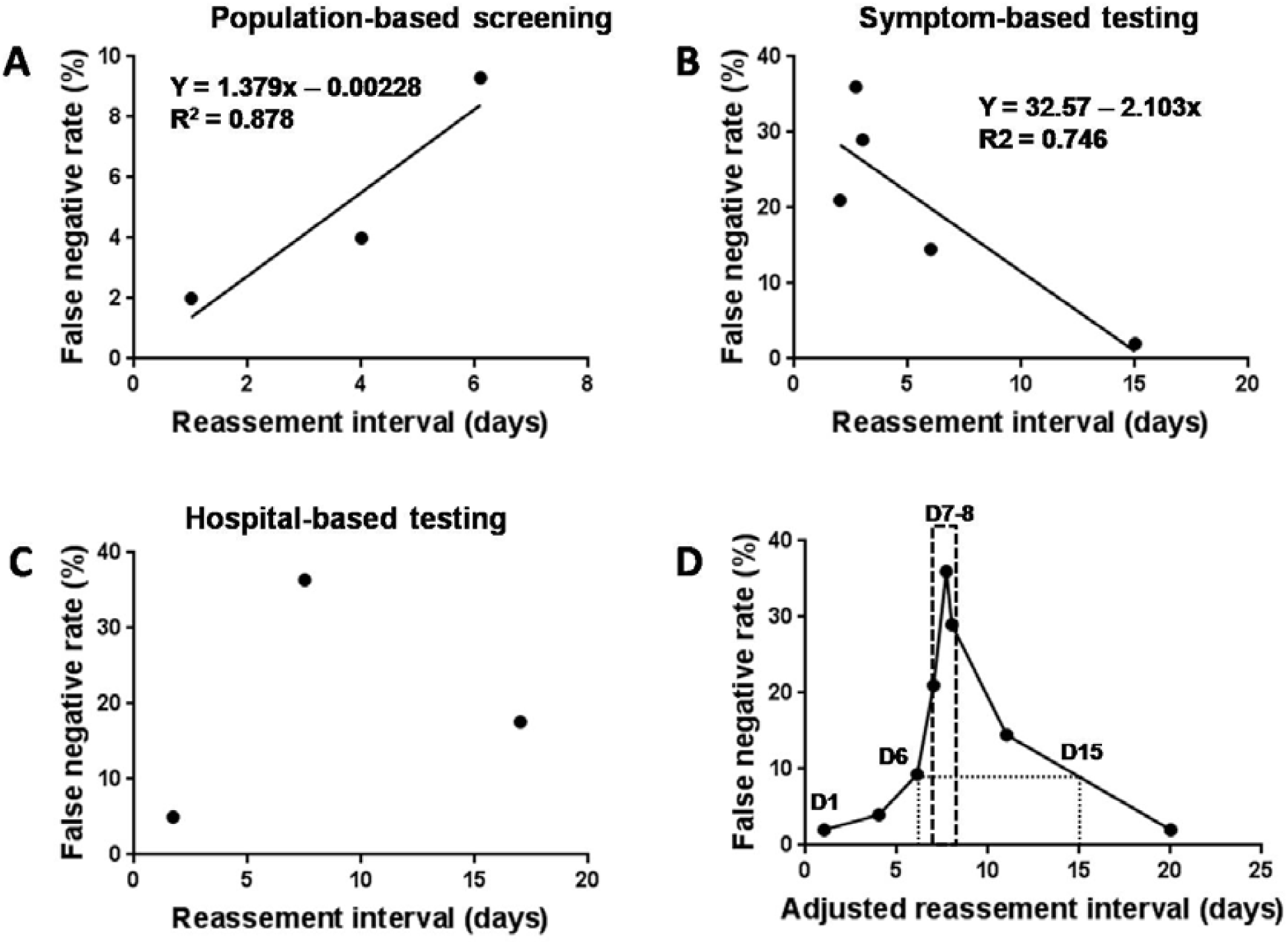
Plot of rate of detection of false negative results in initial RT-PCR against time interval to re-assessment based on repeat RT-PCR or CT scan in (A) population based SARS-COV-2 screening studies, (B) symptom-based testing and (C) hospitalization-based testing. (D) Variation of rate of detection of false negative results in initial RT-PCR against adjusted time interval to reassessment.

PCR could not be accounted for in the adjusted time interval since none of the above studies mentioned that clearly. Data from hospital-based tests could also not be combined since interval between infection or symptom onset and hospitalization is highly variable and context-dependent. The plot indicated that repeat test after 15 days of initial RT-PCR could reduce the identification of false negative results below 10%.

## Discussion

Mutation is an integral part of the evolution of a pathogenic virus. The most challenging consequences of a mutation include increase in infectivity, decrease in susceptibility to neutralizing antibodies due antigenic drift as well as decrease in sensitivity towards detection by existing methods. The SARS-COV-2 enters the host cell by high affinity attachment through hACE2 receptor with a dissociation constant of 4.7 nM. ^27^ Our analysis showed that the N501Y mutation may further enhance this affinity through a stronger inter-molecular H-bonding network. The 501, 498 and 496 residues are located in a relatively flexible region of the RBD between an alpha helical turn and a beta-sheet. Thus the enhanced intramolecular H-bonding involving the 496 and Y501may also provide conformational stability to allow such stronger inter-molecular interactions leading to stronger affinity and increased infectivity associated with emerging variants from UK, South Africa and Brazil harbouring N501Y mutation. However, the fact that this mutation has been there since March 2020, indicates either inadequate sequencing of samples associated with higher infectivity or mechanistic contribution of other mutations that needs to be investigated. Along with increase in hACE2 affinity and viral load, contribution of other mutations in B1.1.7 variant in affecting the pathobiology should be examined to explain the suspected increase in mortality. In addition, recent results indicate that N501Y may itself affect interaction with some antibodies. ^66^

The spike protein RBD also binds to neutralizing antibodies such as P2B-2F6, ^27^ C002, C121, C135 and C144 generated by immune system. ^33^ However, the neutralizing antibodies seem to approach the RBD at a different angle (compared to the hACE2) that does not involve interaction with 501 reside. Earlier study showed that the spike RDB and P2B-2F6 interacts with a dissociation constant of 5.14 nM ^27^ making the neutralizing antibody competitive against hACE2, which is essential to prevent infection. The interaction involves H-bond, salt-bridge as well as Van der Waals interactions involving several residues at the RBD-antibody interface. E484K mutation disrupts the salt bridge and H-bonding interaction with R112 of the heavy chain as well as H-bond with Y34 and N33 of the light chain. The Coulombic repulsion between R112 and K484 also weakens several other intermolecular H-bonds. Complex with C002 shows E484 side chain interacting with R96 (heavy chain) and H35 (light chain), both with positively charged side chains. ^27^ So, similar to that observed in case P2B-2F6, K484 side chain would feel the Coulombic repulsion and disrupt these H-bonds. C121-RBD complex involves H-bond between side chain -OH groups of Y33 and S52 (heavy chain) with same E484 side chain oxygen, ^27^ which has to serve as a donor in one and acceptor in the other H-bond. The C144-RDB complex also shows the E484 side chain oxygen in similar simultaneous donor acceptor role in H-bond with S53 and S56 residue of the heavy chain. ^27^ E484K mutation would disrupt such interactions as well given that the lysine side-chain is two carbons longer and the side chain amine of the lysine is likely to be protonated and hence serve poorly as an acceptor. Thus, E484K mutation would compromise neutralizing activity of several antibodies and ‘escape’ as has been documented. ^35^ On the other hand, C135 neutralizing antibody interacts with N440 side chain of RBD via H-bond network involving D53 and R55 of the heavy chain. ^27^ Their interaction also engages R346 side chain of RBD in H-bond with backbone carbonyls of Y98 (heavy chain) and Y91 (light chain) as well as a pi-cation interaction with the W32 (light chain) aromatic ring. ^27^ This would explain the observed reduction in susceptibility associated with N440K and R346S mutations.^35, 36^ In general, mutations causing significant changes in side chain polarity, charge as well as length or steric bulk of RBD residues that may participate in intermolecular H-bond (E484, K417, K444, N448, N487, Q493, Q498, N440, R346), salt bridge (E484, K417, K444, R346) as well as significant Van der Waals interactions (L452, Y453, A475, V483, F486, F490) are expected to compromise the neutralizing activity. Thus, it was not surprising that earlier studies have found mutations like E484K, K417N as well as L452R, A475V and V483A to result in reduced vulnerability to neutralizing antibodies pesumably due to disruption of H-bond/salt-bridge and Van der Waals interactions, respectively. In fact, an earlier study on in vitro evolution of RBD as well as non-RBD spike protein variants showed that all escape mutants were associated with either change in side chain charge and/or size and hydrophobicity. ^36^ D796H and H655Y mutations in the spike protein, which has also been associated to compromised susceptibility to neutralizing antibodies, ^18, 36^ also involve reversal or removal of side chain charges and are indicative of these residues being involved in H-bond/salt-bridge interaction with antibody. It would be interesting to examine if the L18F mutation found in the B1.1.7 variant contributes to suggested increase in mortality due to disruption of Van der Waals interaction involving the L18 residue. Such mutations significantly affecting spike-antibody interaction could compromise the ability of a person previously infected with the Wuhan strain to neutralize the mutant strain. In case the new variant also harbours a mutation strengthening binding with the competing hACE2 receptor (e.g., N501Y), the neutralizing antibodies could lose decisively leading to re-infection as has been documented for the P1 Manaus strain from Brazil harbouring E484K along with N501Y mutation. ^67^ Co-occurrences and accumulation of these mutations could elevate reinfection risk as well as pose challenge to vaccination and antibody-based therapies.

It is interesting to note that while E484K and other mutations mentioned above could be traced back to March-April, 2020. Also, E484K, K417N and other mutations associated with reduced antibody susceptibility and escape from neutralization are already present in many countries outside Brazil, South Africa or UK. However, re-infections related to them have been documented only recently and still remains modest in number. One of the reasons might have been due to inadequate sequencing and/or detection of the first infection, which may have been asymptomatic. The GISAID database is currently overrepresented by sequences from developing countries. It does not represent the actual geographical distribution of SAR-COV-2 genotypes since many developing countries including India, Brazil, Russia and South Africa has very high number of cases per sequence deposited from those countries. Thus, it is possible that there is a hidden variant load in many of these countries. This is presumably a result of the fact that sequencing is both time-consuming and resource-intensive. However, given that mutations of concern have been reported since March-April, 2020 and are already present in several countries suggest that the possibility of their independent emergence and evolution into strains of concern in different parts of the world is quite real. The difference in mutation profile of two Brazil-related variants identified is also indicative independent branches of evolution existing side-by-side. Novel strains with N501Y has now been detected in Ohio, ^68^ USA, whereas, P681H detected in Nigeria, Δ69-70HV detected in Connecticut and Wisconsin, USA ^70^ and L452R detected in Southern California ^71^ independently. It’s interesting to note that while sample containing the UK B1.1.7 variant were first collected in September 20, 2020, the first set of UK-focused travel restrictions by EU members were imposed two months after that. Several other countries followed that and escalated sequencing of samples from COVID-positive travellers from UK. So, at least for two months, there was possibility of people with the variant travelling across country and continents.

It should be noted that even when the whole world was under lockdown and recent COVID-negative status was mandatory to fly; several people were being detected as COVID-positive upon arrival. It’s not surprising that with the requirement of recent COVID-negative results to fly even now, several travellers from UK and Europe are being detected with the SARS-COV-2 including the mutant strain. For example, India has now detected 150 cases of the UK variant. ^23^ One plausible reason could be that these people picked the infection somewhere on their way to boarding or on board from a fellow traveller. The other possibility is that pre-boarding RT-PCR tests yielded false negative results allowing a COVID-positive person onboard. It should be remembered that this person as well as inoculated fellow travellers, may return a false negative result even at arrival and then spread the strain in the country of destination. For example, the reported B1.1.7 tally in India increased from 58 in January 5 ^72^ to 150 on January 23 ^23^ in spite of ongoing travel restriction and mandatory testing on arrival. Thus, there is every possibility that persons carrying novel strains, such as B.1.1.7 did deliver disseminate them to new destinations during September 20, 2020 and December 20, 2020, when EU countries re-imposed travel restrictions from UK. ^73^ Also, given that travel restrictions have been UK-centric to start with, they may not have prevented spreading of other strains of concern as well as the B1.1.7 strain from a third country.

So, along with escalated sequencing, measures to reduce false negatives results and to track persons with false negative RT-PCR are very important. In case there are no issues with sampling, sample processing issues or RT-PCR primer and protocol, false negatives may arise due to inadequate viral load. A passenger may have low viral load at arrival either due to being infected just during air travel, which would not generally 1-2 days, or because of low dose exposure and better innate immune response. Most of such patients are expected to be asymptomatic as well. A pooled analysis estimated median probability of false negative result to be lowest (20%) at day 8 post-infection, while it is was 66% on day 21 and 100% on day 1. ^39^ Our analysis assuming median incubation period of 5 days indicated that repeat testing after 7-8 days of initial RT-PCR would result in detection of highest number of false negatives in the initial RT-PCR. A recent meta-analysis revised the median incubation period to 7.7 days, ^74^ which would extend the repeat test window to 9.7-10.7 days. So, a repeat test at 7-11 days after the initial RTPCR at arrival along with the current requirement of RT-PCR-negative status immediately before boarding would significantly help to reduce false negative results and reduce transmission. However, as our analysis indicate, reassessment beyond 15 days may significantly compromise the ability to identify initial false negative results.

Given that mutants of concern seem to be quite well-spread around the world and may be evolving further, it would be prudent to warrant repeat assessment for all international travellers. It would be best to keep them in quarantine for two weeks at least till their reassessment results are available. However, it should be noted that it may not be universally acceptable to quarantine all travellers for that long. So, in case the RT-PCR on arrival is negative, which upon repeat test turns out to be false negative, and the traveller is allowed to leave the airport, the variant can spread, particularly if the subject dwells in a densely populated area. Thus, irrespective of initial RT-PCR results, all travellers should be counselled to come under digital surveillance to identify their dwell zone till the initial negative result is confirmed through re-assessment. This would help in monitoring any unusual spike in disease dynamics in these areas and take appropriate actions in case the person is eventually found to harbour a variant of concern through sequencing. It should, however, be noted that some mutations may increase the likelihood for false negative results in RT-PCR. ^75^ In fact, the ΔH69/ΔV70 mutation was found to affect the result of diagnostic RTPCR tests with S target gene. ^10^ Thus, the sensitivity of the strain of interest should be thoroughly monitored and multiplexed PCR protocols should be adopted. ^76^ Development of strain-specific primers ^77^ and RT-PCR protocols should be taken up expeditiously so that surveillance for strains interest can be improved even without mandatory sequencing, which is time-consuming, expensive and requires specific expertise.

In view of the fact that large number asymptomatic travellers with false negative tests may have already escaped detection under the current protocol of test at arrival, travellers from countries reporting emerging variants dating back to, at least, September, 2020 should be catalogued and their dwell zones should be mapped. These areas should be monitored and the temporal evolution of COVID case load, severity and mortality maps should be compared with dwell zone maps to examine any correlation, which should warrant escalated testing and sequencing effort.

Mutation and antigenic drift is an integral part of the evolution of a pathogenic virus. While it may be interesting that the emerging variant B1.1.7 has acquired fourteen mutations over the Wuhan strain, we should have seen it coming. It should not be any surprise for a virus that apparently could accumulate several mutations, including an insertion at the functionally crucial S1/S2 cleavage site, in it’s spike protein alone over it’s bat variant to infect humans. ^78^ In fact, it’s ability to swiftly mutate and transmit back and forth between human and animals is manifest from the SARS-COV-2 mutations including N501T, F486L and Y453F in the spike protein RBD detected in connection with infections in mink farms. ^79^ The RBD mutations like Y453F was also found in cats whereas N501T was found in ferrets and mice were found to have K417N, Q493H/K, Q498H as well as N501Y mutations. ^80^ A recent study showed presence of N501T in a human variant along with Q493K that went undetected and has been estimated to have evolved in Italy in early August, 2020. ^81^ Our analysis showed N501T to be first reported in March, 2020 and now present in 12 countries including Italy with counts increasing steadily. Our analysis also showed that the N501Y as well as E484K, K417N and other mutations that have been shown to be associated with reduced susceptibility to antibodies have been detected as early as in March-April, 2020. These mutations are already present in several countries across the world besides UK, Brazil and South Africa. Double SNVs such as N501Y/K484K or N501Y/K417N found in the Brazilian and South African variants were detected as early as in October, 2020. These are indicative of the fact that these mutations have been accumulating in SARS-COV-2 and circulating around for quite some time. They only raised alarm and got identified since their cluster stood out against the background of a decreasing number of cases, probably, associated with an earlier and relatively less infectious strain. It is interesting to note that out of five emerging strains of concern; only the one reported from South Africa had D614G mutation in spite of the fact that D614G has been a super-dominant mutation across the world since March, 2020. It may be indicative of independent and isolated evolution of other four variants which may precede that of the South African variant. The other possibility is a very rapid evolution one of the very few non-D614G strain bypassing transmissional advantage of the D614G strain. ^15,16^ This would be possible if it evolves within the same host. It has been shown that the virus can evolve within immunocompromised patients ^82^. Deletion in the Y145 region has been found in immunocompromised patients and it was speculated that the B1.1.7 may have initially evolved in such patients. ^83^ This indicates to possibility of emergence of distinct variants without necessary linkage with dominant strains around. It is important to note immunocompromised patients also show prolonged shedding.^82, 84^ Apart from patients under treatment for conditions like cancer, organ transplant, autoimmune diseases, SARS-COV-2 patients also receive immunosuppressive agents like corticosteroids. Thus, hospitals with such patients are potentials hot zones for SARS-COV-2 evolution and should be under rigorous surveillance and escalated sequencing. In fact, in January 2021, a cluster of 35 new infections has been detected in a hospital in Germany ^85^ potentially belonging to a new strain that remains to be sequenced. Given the agility of SARS-COV-2 to evolve to and fro between animals and humans, animals farms should also be under surveillance to detect and contain emergence of any new variant at the earliest.

In conjunction with existing knowledge about other viruses like influenza, ^86, 87^ we should also expect further mutations and antigenic drift during SARS-COV-2 evolution across the world. Mutations with simple increase in infectivity without any bearing on immune response are likely to increase overall number of casualties and may also increase in mortality among old and vulnerable. However, ‘escape mutations’, like E484K, ^35-37^ leading to reduced efficacy of neutralizing antibodies are more likely to increase disease severity, duration and poor outcome even among healthier patients. In addition, reduced efficacy of antibodies would increase risk of re-infection along with possibility of antibody-dependent enhancement in previously infected or vaccinated individuals. Thus, any significant spike in hospitalization and disease severity or duration among young and healthier individuals should be examined as a possible sign of emergence of a variant of concern. Since younger people are more active and many of them may still be asymptomatic or mildly symptomatic, they can spread the variant fast, especially, in areas with high population density. So, surveillance mechanism should allow examination of demographic distribution of evolving scenario. For any suspected case, similar to that in case of travellers, repeat RT-PCR test should be performed 7-11 days after the initial test.

While an earlier version of this work with similar recommendations was submitted to the Lancet (January 4, 2021) for consideration as a correspondence article, WHO expert group met on January 12 to give a series of recommendations on increasing surveillance, testing, sequencing and calling for evidence-based strategy to mitigate challenges faced by emerging variants. ^88^ Subsequently, a correspondence article on action plan for pan-EU defence against emerging variants was published. Elaborate recommendations included increased surveillance, sequencing and specific mention of repeat test 7-10 days after travel along with RT-PCR negative status within 24 hours prior to travel and 10 days strict quarantine. ^89^ Our analysis showed the threat of emerging variants are more widespread than currently assumed and may not be contained with prospective travel restrictions and testing alone. Some of these mutants may already have crossed borders in absence of appropriate testing or sequencing protocols and many more may independently emerge in different parts of the world, particularly, in hospitals, animal farms and densely populated areas. It is noted with concern that a recent analysis revealed that N501Y, E484K and K417N mutants show significant resistance to antibodies elicited by mRNA vaccines that are currently being used for immunization. ^66^ Analysis and specific suggestions made in this article should help to take a more calibrated approach towards surveillance, socio-economic activities, vaccination and antibody-based therapies to deal with the pandemic in real time.

## Data Availability

All data used in this study are publicly available through Protein Data Bank, GESS database and published articles.

https://wan-bioinfo.shinyapps.io/GESS/

https://www.rcsb.org/

## Summary

This study revealed the structural basis of increased infectivity and reduced antibody sensitivity associated with emerging variants. It showed that several mutations associated with such effects emerged very early during the pandemic and may have been evolving independently. It analyzed the available data to arrive at the following specific recommendations to deal with challenges of emerging variants. (1) At least one repeat RTPCR testing of travellers or contacts or any suspected individual 7-11 days after the initial RT-PCR test at arrival or referral, (2) digital surveillance on dwell zone of travellers for COVID-19 dynamics in case mandatory two weeks quarantine cannot be imposed, (3) retrospective cataloguing of international travellers from countries reporting emerging variants and analysis of COVID-19 dynamics in their dwell zones, (4) escalation in sequencing and development of variant-specific and - sensitive diagnostic method, (5) surveillance of hospital and animal farms, (6) monitoring of any unusual spike in disease burden, severity and outcome among healthy, young individuals, (7) monitoring of any spike in re-infection or adverse health problems in vaccinated individuals.

## Supplementary Material

Supplementary figures and legends can be found in supplementary material.

## Acknowledgement

Authors are supported by funding from the Department of Atomic Energy, Government of India.

## Supplementary Figure Legends

**Figure S1.**
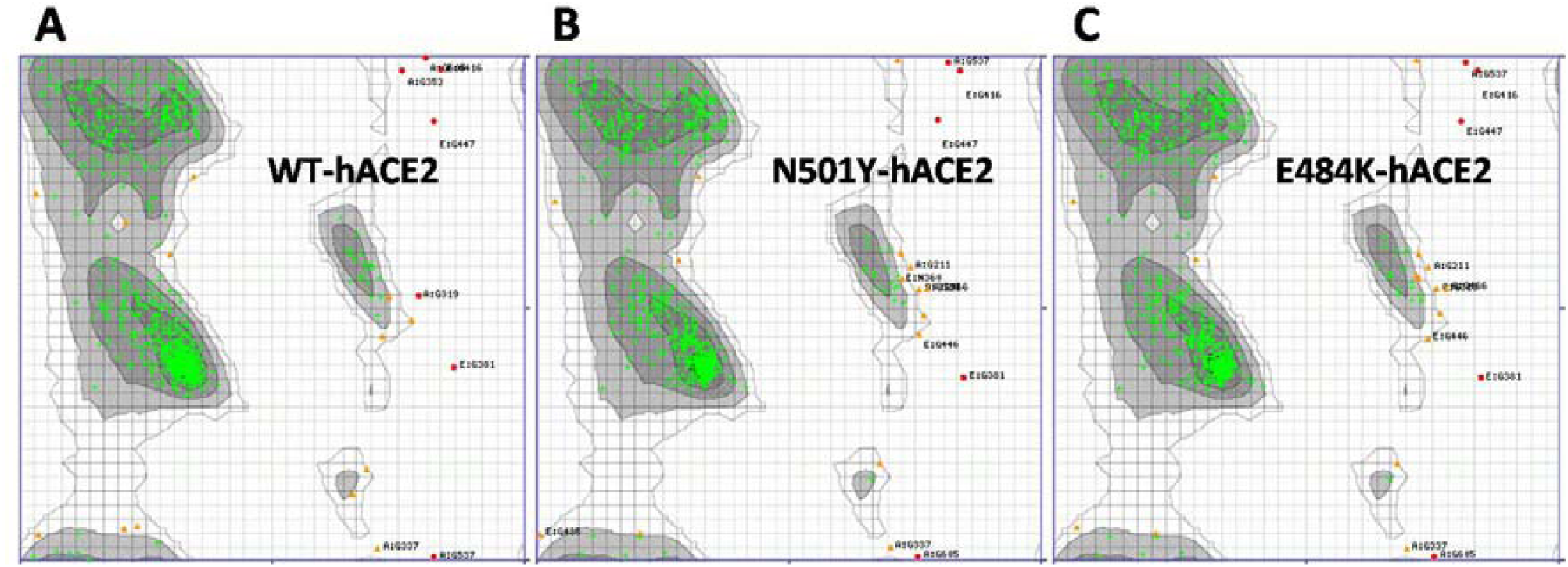
Analysis of dihedral angle of amino acid residues and their position in the Ramachandran plot for (A) Wuhan strain (PDB: 6M0J), (B) N501Y mutant and (C) E484K mutant of SARS-COV-2 spike protein receptor binding domain bound to the human ACE2 receptor. PDB files for mutant complexes (see supplementary material) generated via MutaBind2 server. Green, orange and red dots indicates residues in highly preferred, preferred and questionable conformation. Only glycine residues were found to be in questionable conformation.

**Figure S2.**
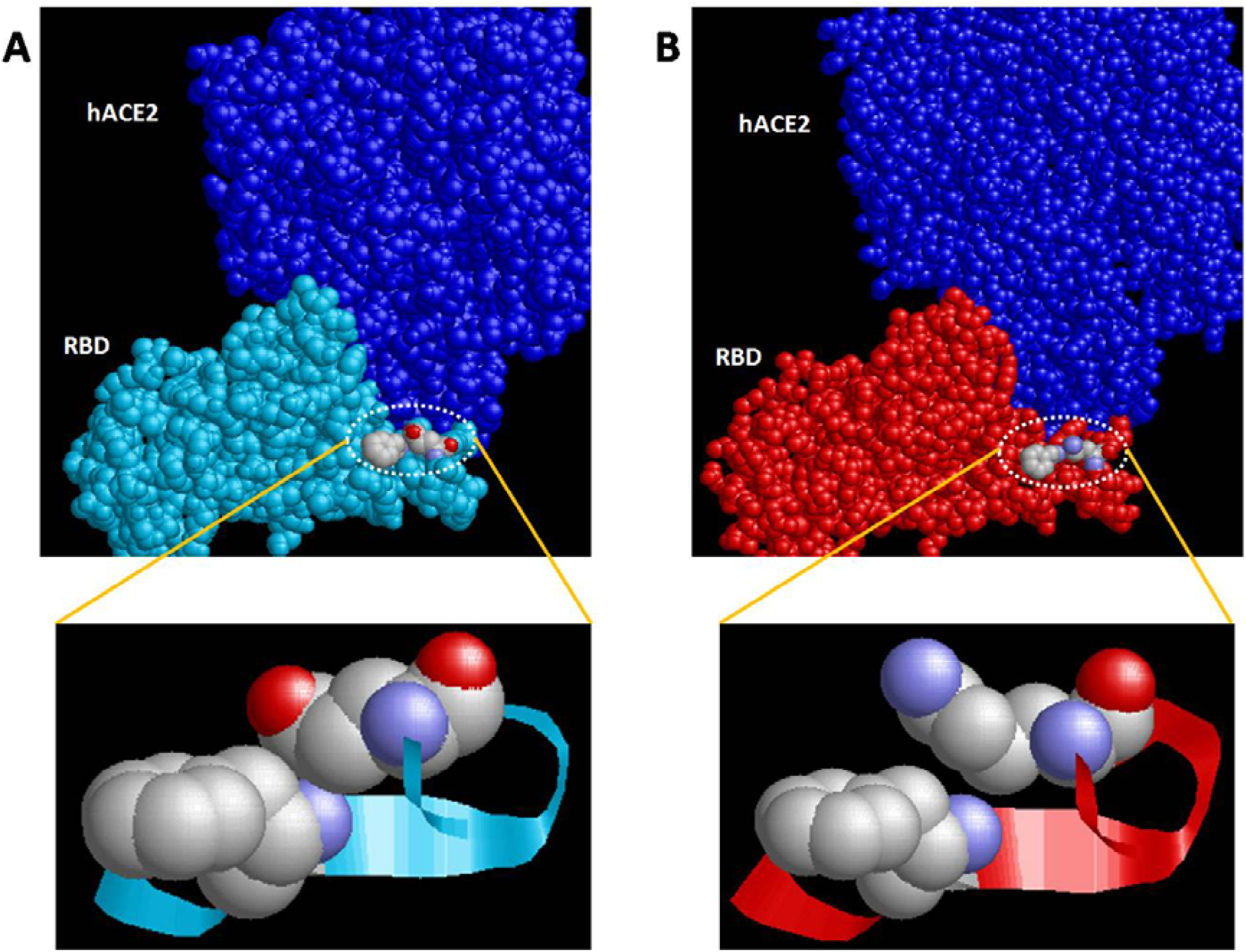
Spacefill models showing surface exposed (A) E484 residue of the spike RBD domain of the Wuhan strain (shown in cyan) and (B) K484 residue of the E484K mutant (in red) of SARS-COV-2 along with F490 residue in complex with the human ACE2 receptor (in blue). The F490 residue below the

**Figure S3.**
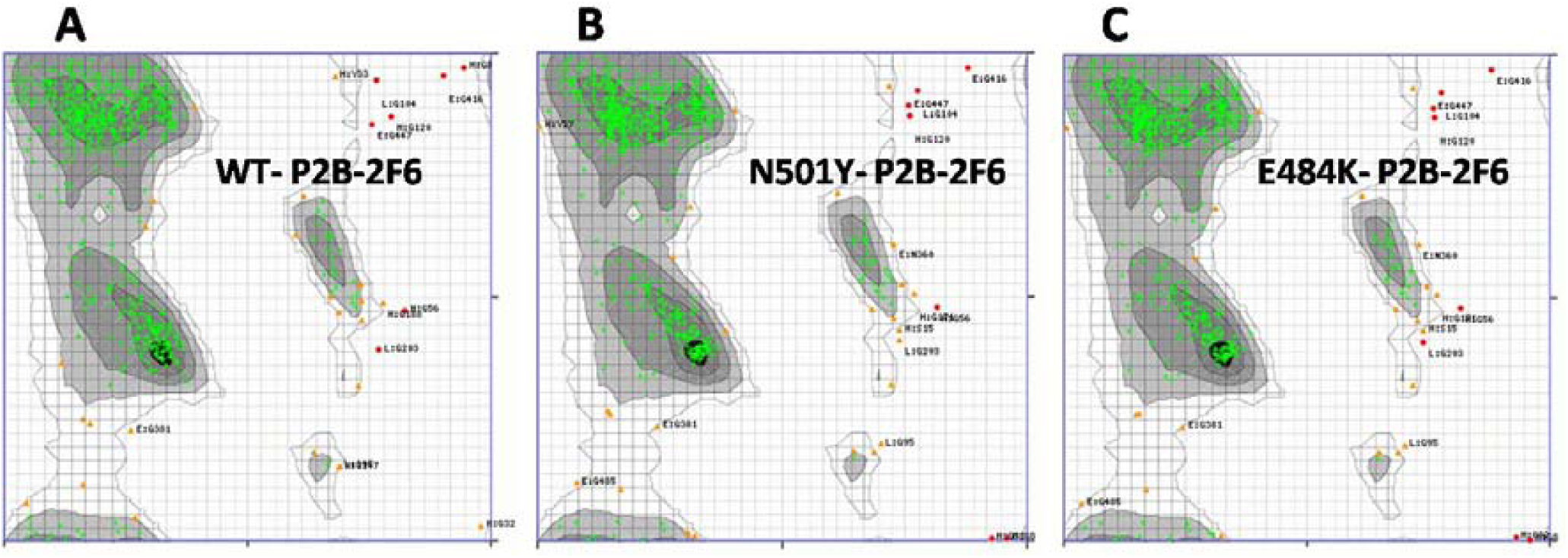
Analysis of dihedral angle of amino acid residues and their position in the Ramachandran plot for (A) Wuhan strain (PDB: 7BWJ), (B) N501Y mutant and (C) E484K mutant of SARS-COV-2 spike protein receptor binding domain in complex with P2B-2F6 neutralizing antibody. PDB files for mutant complexes (see supplementary material) generated via MutaBind2 server. Green, orange and red dots indicates residues in highly preferred, preferred and questionable conformation. Only glycine residues were found to be in questionable conformation.

**Figure S4.**
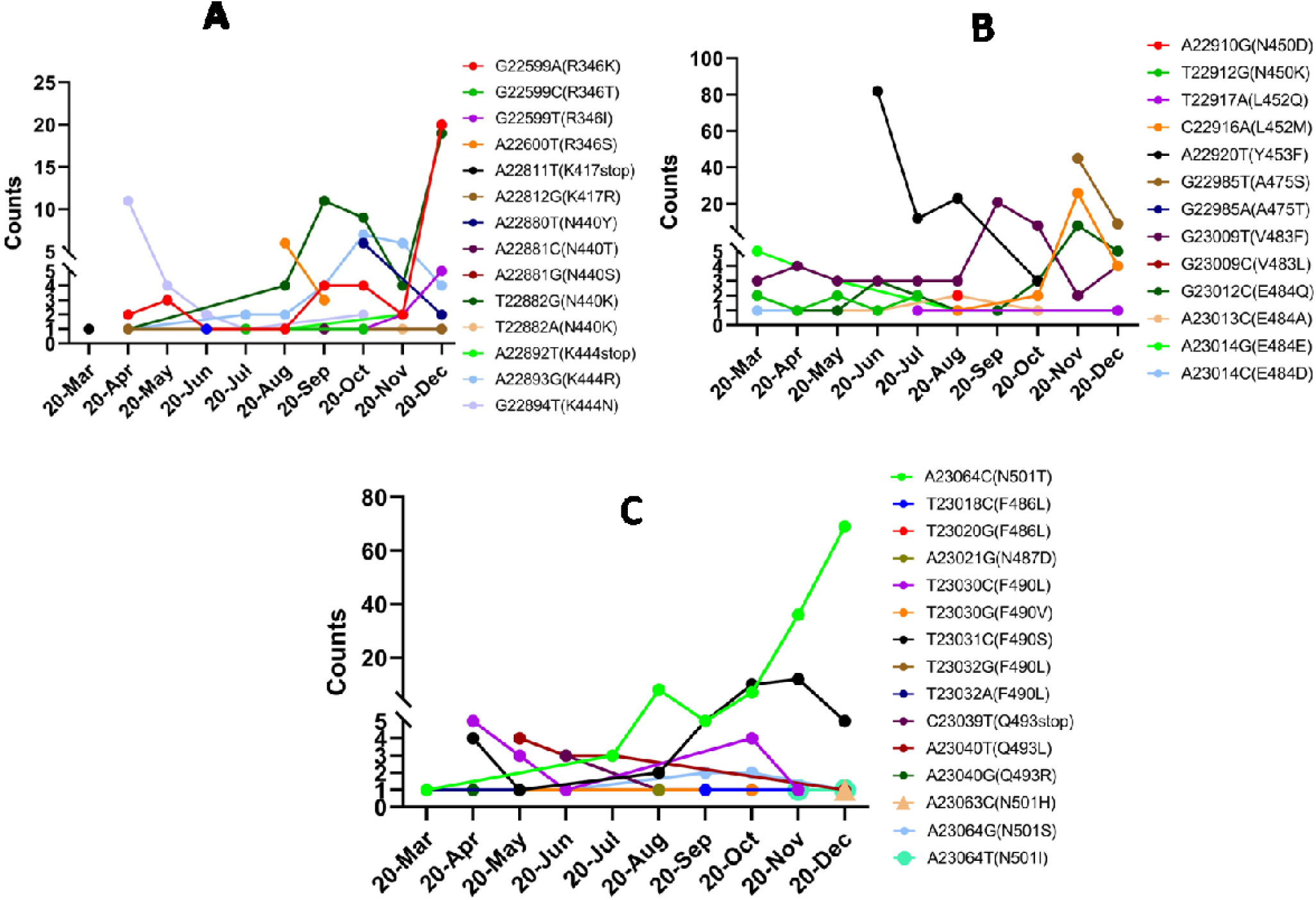
Monthly variations in SNVs detected in (A) 346, 417, 444, (B) 448, 450, 452, 453, 475, 483, 484, (C) 486, 487, 490 and 493 positions of spike protein RBD shown to be involved in interaction with neutralizing antibodies.

**Figure S5.**
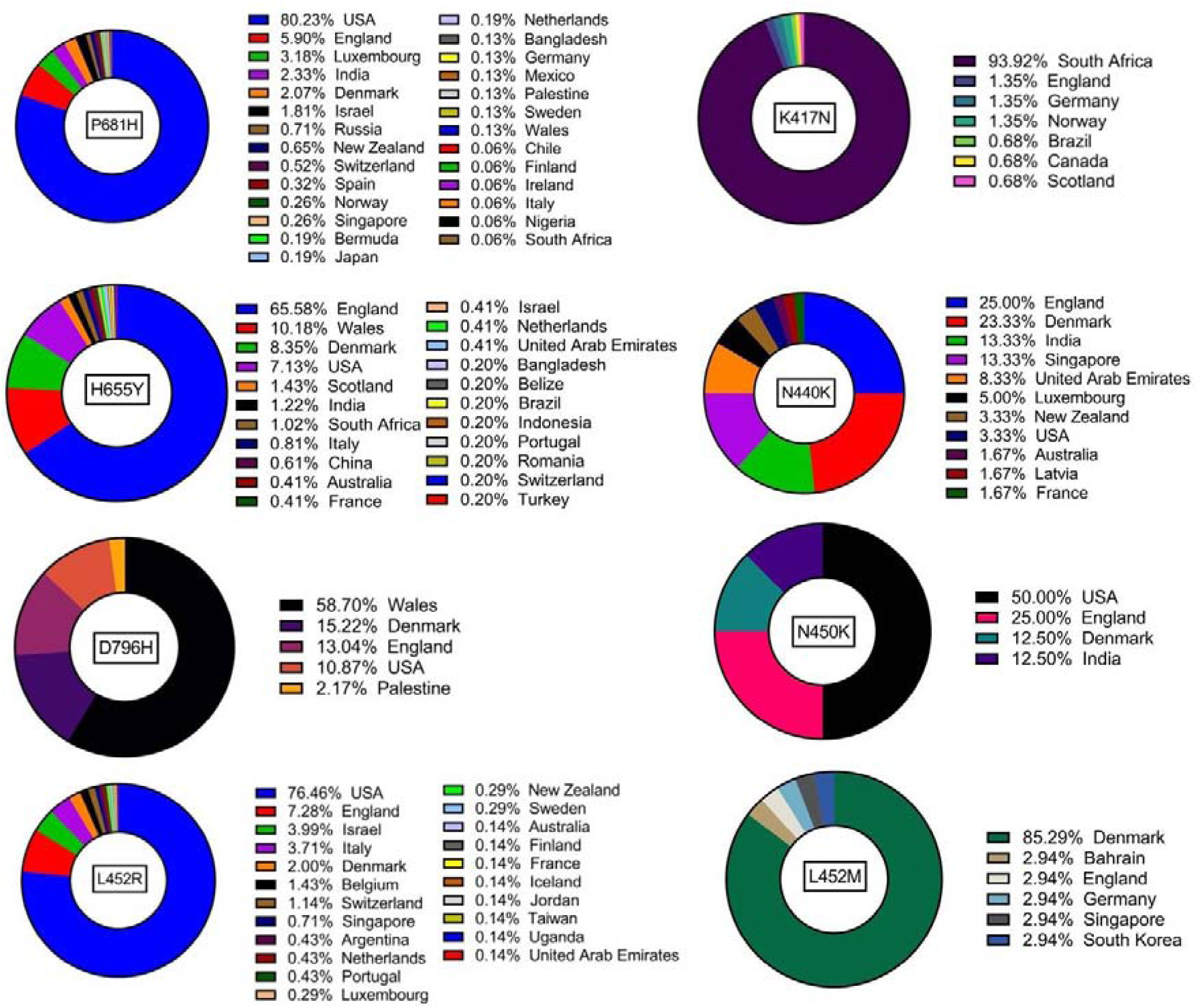
Figure S5. Country-wise relative abundance of P681H, K417N, H655Y, N440K, D796H, N450K, L452R and L452M mutations as reported in GESS database.

**Figure S6.**
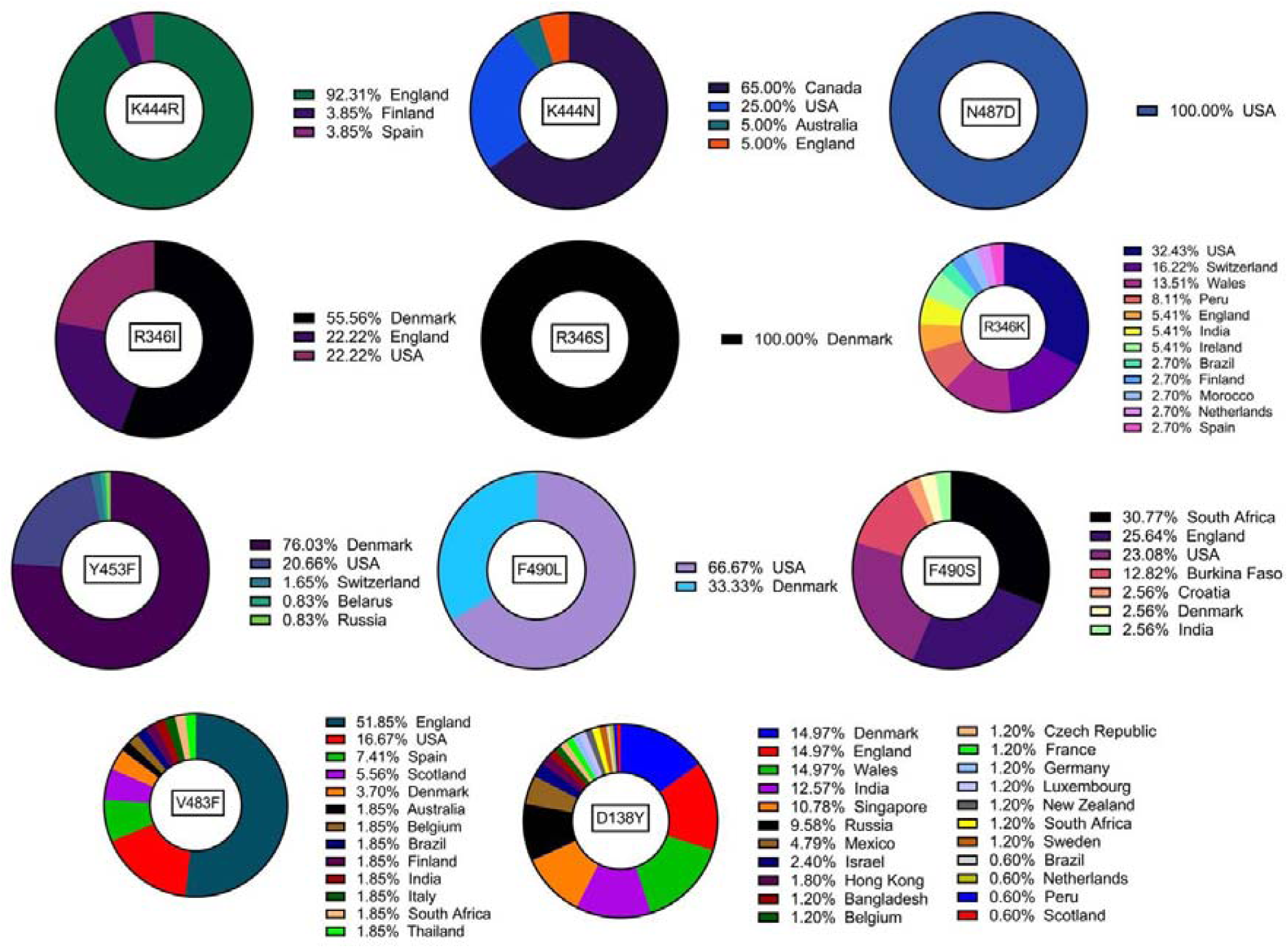
Country-wise relative abundance of K444R, K444N, N487D, R346I, R346S, R346K, Y453F, F490L, F490S, V483F, D138Y mutations as reported in GESS database.

